# Understanding the acceptability of digital diabetes self-management tools among non-users: A qualitative study of personal, social and healthcare contexts

**DOI:** 10.64898/2026.09.29.26364317

**Authors:** Rhiannon E. Hawkes, Hibbah Osei-Kwasi, Daniel Taylor, Dennis Ashpole, Cerys E. F. Reiger, Daniella M. A. Amutike, Mats Baxter, Alex Bickerton, Scott G. Cunningham, Deborah J. Wake, Tony Willis, Martin K. Rutter, Joanna Brooks

## Abstract

**Background:** Diabetes poses a major global health and economic burden, and effective self-management is essential for preventing complications. The National Health Service in England has recently announced plans to shift from analogue to digital services in their 10-year plan. However, digital health technology may also risk widening existing inequalities. MyWay Diabetes is a digital self-management intervention implemented in three regions across England. Understanding the acceptability of online and digital tools to support diabetes self-management, particularly among non-users and underserved groups, is essential to ensure equitable engagement and implementation.

**Objectives:** To (1) explore barriers to using online and digital support across diverse demographic groups with type 1 and type 2 diabetes who had not used the MyWay Diabetes platform, and (2) identify factors that could improve uptake and continued engagement.

**Design:** Cross-sectional qualitative study.

**Methods:** We conducted six focus groups with n=35 people living with diabetes (27F, 8M; 40% Asian British Pakistani) across three regions in England. The Theoretical Framework of Acceptability (TFA) informed data collection and analysis. Data were transcribed and analysed thematically using template analysis, mapped onto constructs of the TFA.

**Results:** Digital engagement in diabetes self-management was shaped by intersecting personal, social, and healthcare system factors. Participants’ confidence with navigating digital environments was constrained by the emotional and practical burden of living with diabetes. Social and structural barriers, including cultural mismatch, accessibility challenges, financial constraints, and reliance on family further limited meaningful use of digital tools. Engagement was strongly influenced by trust in healthcare professionals and services; digital tools were considered acceptable when they complemented person-centred care, but resisted when perceived as replacing personalised, face-to-face support.

**Conclusions:** Digital support for diabetes self-management is more likely to be acceptable when it aligns with people’s capabilities, social circumstances, and trusted healthcare relationships. Therefore, to prevent widening inequalities, digital tools should be accessible, culturally relevant, and embedded within supportive, person-centred models of care.

**Lived Experience or Public Contribution:** Public contributors with lived experience of diabetes (3F, 3M) reviewed participant materials and pseudonymised focus group transcripts, providing feedback on recruitment, data collection, and interpretation. Their insights informed analysis and theme development, ensuring a lived-experience perspective. Our public co-applicant is a co-author on this manuscript.

## Introduction

Diabetes is a major global and UK public health challenge with substantial healthcare costs and preventable complications, making effective self-management essential [1–5]. Diabetes self-management education and support (DSMES) programmes play a central role in enabling people with diabetes to manage their condition effectively and are associated with improved clinical and psychological outcomes. Despite these benefits, uptake of face-to-face provision remains low [6,7].

Digital interventions offer promising solutions to some of the challenges associated with traditional face-to-face programmes [8–10]. Digital approaches to diabetes self-management have been found to be effective, well-received by users, and supportive of autonomy by those who use them [11,12]. However, technology in healthcare may also risk widening existing inequalities [10], and it is therefore important to explore access to diabetes care when digital DSMES and technologies are integrated into care pathways [13]. A scoping review led by Public Health Wales found that people with greater health needs and language barriers still struggle to use digital health services and technologies, mainly due to limited access, low digital literacy and services being poorly adapted to diverse needs [14]. This is particularly important in the context of the NHS 10-year plan in England, which aims to reform the health service by shifting from analogue to digital services [15].

MyWay Diabetes is a digital life-long self-management platform/tool that provides DSMES as part of its core offering, available via web and mobile applications. It has been implemented in Scotland (as ‘My Diabetes My Way’ [16], developed by the University of Dundee) since 2008, and in several regions in England since 2018-2020, including Somerset (‘MyWay Diabetes’), Greater Manchester (‘Diabetes MyWay’), and North West London (‘KnowDiabetes’). As with other healthcare interventions, understanding acceptability as well as effectiveness is essential for equitable implementation [17]. Acceptability assessments can yield critical insights into usability, accessibility, and the likelihood of uptake. This is particularly pertinent, given that uptake of ‘My Diabetes My Way’ in Scotland is historically lower in people from ethnic minorities and those who live in more deprived areas [18,19] and barriers such as limited digital literacy identified in user surveys of MyWay Diabetes [20].

While recent qualitative work has explored the acceptability of MyWay Diabetes among clinicians and current users [21], less is known about the perspectives of people who do not use the platform. Diabetes prevalence, care quality, and outcomes are disproportionately worse among ethnic minority and socioeconomically disadvantaged groups [22]. Identifying factors that may hinder engagement with digital technologies is essential to ensure digital DSMES interventions are accessible and acceptable to diverse population groups. This has been highlighted by evaluations of other nationally implemented digital diabetes programmes in England [23,24].

This qualitative study was nested within a larger mixed-methods evaluation of the MyWay Diabetes platform in Greater Manchester, Somerset and North West London, UK [25]. We aimed to (1) explore barriers to using online and digital support across a diverse cohort of people with type 1 and type 2 diabetes who had not used the MyWay Diabetes platform, and (2) identify factors that could improve uptake and continued engagement. By examining perspectives from people of different ages, ethnicities, socioeconomic backgrounds, and geographical regions, this study sought to identify modifiable factors that could enhance the acceptability of MyWay Diabetes and inform the wider design and implementation of digital DSMES interventions.

## Methods

Methods are reported in accordance with journal article reporting standards for primary qualitative research in psychology (JARS-Qual; [26]).

### Study Design

This was a cross-sectional qualitative study, using focus groups with adults living with diabetes to discuss barriers to using online and digital services to manage their condition.

### Participants

Participants were adults (≥18 years) with type 1 or type 2 diabetes, living in Greater Manchester, Somerset or North West London, who were not registered on the MyWay Diabetes platform and were able to provide informed consent.

### Sampling and Procedures

A community-based sampling approach was taken. We worked with local community contacts to identify and recruit people from ethnic minority communities and socioeconomically deprived backgrounds, who are disproportionately affected by diabetes, at greater risk of digital exclusion, and less likely to participate in research. We developed relationships with various local community groups via (1) online meetings with the community leaders/trusted advocates, and (2) via engaging with in-person health events organised by community leads in their local community. Contacts distributed study information to eligible participants and in some cases supported recruitment and data collection. See more detail on recruitment processes in Supplementary Material 1.

Focus groups took place between September 2025 and June 2026 and facilitation was shared between REH, JB, CEFR and DMAA. Five focus groups were conducted in person at community venues, and one focus group was conducted online via Microsoft Teams. Participants provided full consent prior to the group discussion taking place, and they completed some optional demographic questions (age, gender, ethnicity, postcode and diabetes duration). Postcodes were only used to extract English Index of Multiple Deprivation scores [27]. Discussions were either audio-recorded using an encrypted dictaphone and transcribed using a third-party transcription company who had signed confidentiality agreements, or were recorded and transcribed using Microsoft Teams (see Supplementary Material 1).

Focus group discussions lasted a median duration of 65 minutes, and field notes were made following each discussion. After each focus group had taken place, participants were provided with a debrief sheet, with signposting to relevant self-management and digital literacy skills resources (e.g., ‘Learn My Way’ platform [28]; Good Things Foundation, [29]).

### Materials

A flexible topic guide to investigate ‘prospective acceptability’ of online and digital support for managing diabetes was informed by the Theoretical Framework of Acceptability (TFA [30]). The TFA is a conceptual model outlining key components that influence how individuals perceive and accept a healthcare intervention. The framework identifies seven component constructs of acceptability: affective attitude, burden, perceived effectiveness, ethicality, intervention coherence, opportunity costs, and self-efficacy [30]. This framework has been used to assess acceptability of digital and mobile interventions [30,31], including digital interventions for type 2 diabetes [32], and acceptability of the pilot rollout of MyWay Diabetes in Greater Manchester in 2020 [33,34]. The topic guide explored perceptions of online and digital support, barriers to uptake, and potential adaptations to improve accessibility and equity (see Supplementary Material 2).

### Data Analysis

Data were analysed thematically using template analysis [35]. Initial data coding was undertaken by REH, using an inductive, data-driven approach to ensure that non-theory delineated factors were not overlooked, and nuance and context were not lost. An initial coding template was then developed based on the TFA constructs plus inductive codes identified during initial data coding and iteratively refined through application to a subset of the data (three focus group transcripts). As higher-order TFA constructs recurred across multiple parts of participant accounts (for example, participants discussed “affective attitude” in relation to their management of diabetes, use of digital tools and wider healthcare structures) an updated template focused on inductive codes, with deductive mapping to TFA constructs conducted separately and summarised in tabular format.

The updated template was iteratively refined through application to further transcripts until it adequately represented all data. A final version template was applied to the full dataset. Established quality-checking procedures were used, including an appropriate audit trail, critical scrutiny, regular discussion and constant comparison of coding between analysts (REH and JB). NVivo software (version 15) supported data management.

### Patient and Public Involvement and Engagement

Patient and public involvement and engagement (PPIE) was embedded throughout the study with support from Vocal [36], a not-for-profit organisation specialising in PPIE in health research. PPIE is reported using GRIPP2 short form guidance [37] in Supplementary Material 3.

Public contributors (n=3 female, n=3 male; purposively recruited across the three regions including users, non-users and past users of MyWay Diabetes) helped refine the participant-facing materials (e.g., study advert), commented on recruitment and data collection processes, and supported interpretation of findings. Contributors were provided with pseudonymised raw focus group transcripts and gave their initial impressions of the discussions, and highlighted aspects of the discussions that resonated with them and/or warranted further exploration (see Supplementary Material 3). The PPIE group also created a separate recommendations report based on their takeaways from the transcripts and their own experiences. The researchers incorporated this scrutiny throughout the analysis and theme development, to ensure the analysis benefited from a lived experience perspective.

### Researcher Positioning

The lead author (REH) is an experienced health psychology researcher with training in qualitative methods and >8 years’ experience in diabetes prevention and self-management research. Researchers CEFR and DMAA conducted some of the focus groups and have training in health psychology and qualitative methods. Researcher JB is a highly experienced qualitative researcher who co-developed the template analysis method [35] with experience of conducting qualitative research across a range of health conditions, including diabetes. HOK provided advisory support for data collection and analysis, drawing on her expertise in qualitative research with underserved and ethnically diverse populations. The wider multidisciplinary research team consisted of consultant diabetologists, digital health researchers, and public involvement specialists. The MyWay Digital Health team were not involved in the data collection or analysis which were undertaken independently. They contributed to the interpretation of the findings by drawing on results of a previous evaluation of ‘My Diabetes My Way’ conducted by the University of Dundee in Scotland.

### Ethical Considerations

This study was reviewed and approved by The University of Manchester Proportionate Research Ethics Committee (ref: 2025-23064-42006). Full consent was obtained from all participants prior to participation. Those who participated could opt to receive a voucher as a ‘thank you’ for taking part. Data presented are pseudonymised.

## Results

We conducted six focus groups with n=35 people with diabetes. Participants comprised more females (n=27, 77%) but varied in ethnicity, age and duration of diabetes. We spoke with n=6 (17%) people with type 1 diabetes and n=29 (83%) people with type 2 diabetes (Table 1).

**Table 1.**
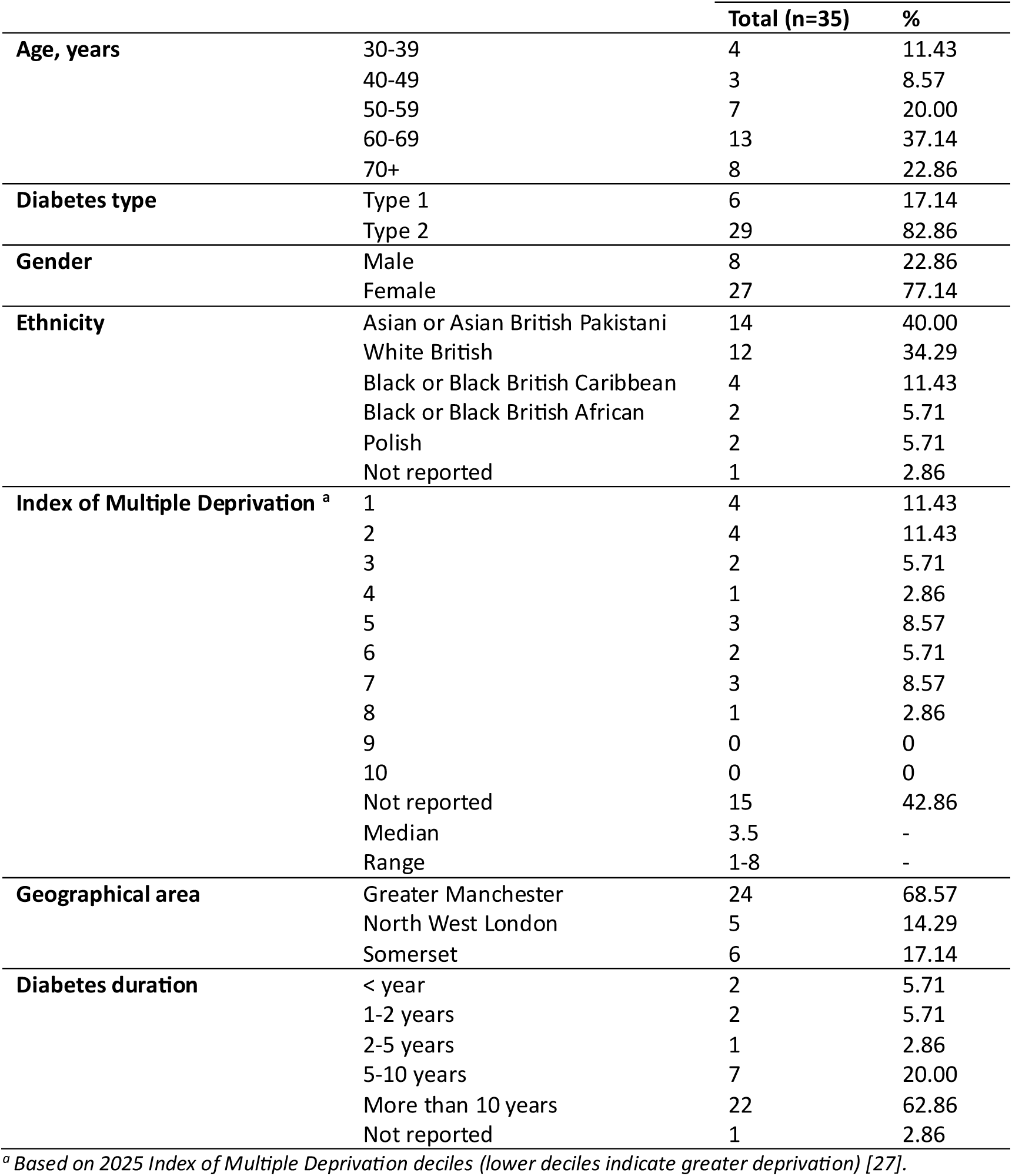
Participant demographic and clinical characteristics.

|  |  | **Total (n=35)** | **%** |
| --- | --- | --- | --- |
| **Age, years** | 30-39 | 4 | 11.43 |
|  | 40-49 | 3 | 8.57 |
|  | 50-59 | 7 | 20.00 |
|  | 60-69 | 13 | 37.14 |
|  | 70+ | 8 | 22.86 |
| **Diabetes type** | Type 1 | 6 | 17.14 |
|  | Type 2 | 29 | 82.86 |
| **Gender** | Male | 8 | 22.86 |
|  | Female | 27 | 77.14 |
| **Ethnicity** | Asian or Asian British Pakistani | 14 | 40.00 |
|  | White British | 12 | 34.29 |
|  | Black or Black British Caribbean | 4 | 11.43 |
|  | Black or Black British African | 2 | 5.71 |
|  | Polish | 2 | 5.71 |
|  | Not reported | 1 | 2.86 |
| **Index of Multiple Deprivation <sup>a</sup>** | 1 | 4 | 11.43 |
|  | 2 | 4 | 11.43 |
|  | 3 | 2 | 5.71 |
|  | 4 | 1 | 2.86 |
|  | 5 | 3 | 8.57 |
|  | 6 | 2 | 5.71 |
|  | 7 | 3 | 8.57 |
|  | 8 | 1 | 2.86 |
|  | 9 | 0 | 0 |
|  | 10 | 0 | 0 |
|  | Not reported | 15 | 42.86 |
|  | Median | 3.5 | - |
|  | Range | 1-8 | - |
| **Geographical area** | Greater Manchester | 24 | 68.57 |
|  | North West London | 5 | 14.29 |
|  | Somerset | 6 | 17.14 |
| **Diabetes duration** | < year | 2 | 5.71 |
|  | 1-2 years | 2 | 5.71 |
|  | 2-5 years | 1 | 2.86 |
|  | 5-10 years | 7 | 20.00 |
|  | More than 10 years | 22 | 62.86 |
|  | Not reported | 1 | 2.86 |

<sup>a</sup> Based on 2025 Index of Multiple Deprivation deciles (lower deciles indicate greater deprivation) [27].

Three themes are presented: (1) ‘*Diabetes self-management and navigating digital environments,*’ (2) ‘*The social conditions of digital self-management,*’ and (3) ‘*Healthcare structures shaping digital engagement*.’ Engagement with online and digital support was not simply a matter of providing tools; rather, it was shaped by the cumulative burdens of self-management (theme 1), the social networks that support daily management (theme 2), and trust in the healthcare system (theme 3).

### Theme 1: Diabetes self-management and navigating digital environments

Participants’ accounts indicated that engagement with digital health tools was shaped not only by attitudes toward technology but also by the ongoing emotional, cognitive, and practical burden of living with diabetes. Two parallel challenges ran through their narratives: the daily burden of managing diabetes, and the uncertainty of navigating digital environments. These challenges often intersected, meaning that digital tools were perceived as potentially beneficial but also adding to the burden of managing diabetes within an already demanding self-management landscape.

Across the focus groups, participants described diabetes management as a continuous process requiring sustained vigilance, behavioural change, and emotional effort. For some, these challenges were intensified by cultural norms and expectations. Participants from Black and African communities, for example, spoke about the shame and taboo surrounding illness and diabetes (“*I think it’s because we’re Black that we feel ashamed of something that’s happened to us,*” **Participant 1, focus group 6**). For many, dietary adjustments were a central and ongoing struggle and participants described the effort involved in changing long-established dietary habits:

> **Participant 1:** *There was a time when I used to put sugar, obviously, like one, one and a half in tea, but I stopped all that…I’ve stopped everything. I’ve got loads of sugar at home, but I do not use it in the tea*.
>
> **Participant 6:** *What about sweetener?*
>
> **Participant 1:** *No sweetener, I stopped all that. I used to have it a year ago but I’ve stopped it, […] like I said, it’s like willpower, isn’t it? I stopped adding it to milk as well. You can do it. I’ve cut sugar*. **Focus group 2**

For those with type 1 diabetes, this burden was described as particularly acute, with participants emphasising the abrupt and life-altering nature of their diagnosis:

> *Yeah, I’ve had it [type 1 diabetes] sort of like what, 5 and a half years now, and never having a problem with food at all, not having to really, being really lucky, not having to watch what I eat, all of a sudden it was like my life’s just changed overnight. Yeah, so I still struggle with my sugar levels*. **Participant 3, focus group 3**

Although some participants reported feeling confident searching for information online in relation to their diabetes self-management (e.g., using Google or YouTube), conflicting information across sources reduced trust and caused further confusion (“*But sometimes, you know, the websites and the information can be different as well,*” **Participant 3, focus group 2**).

Even when motivated to manage their condition well, many participants lacked confidence in their digital skills. Several described themselves as “*not a digital person*” (**Participant 3, focus group 3**), described apps as “*frightening*” (**Participant 3, focus group 6**), and expressed uncertainty about how to judge the reliability of online information and fears of “*clicking something else by mistake*” (**Participant 4, focus group 5**):

> *I can hardly do anything. I try to get my son to help me, and he confuses me more, but I am very, very nervous at technology. If it’s to do with diabetes, I’m really scared, I’ll be honest with you. I’ve got no confidence with it*. **Participant 3, focus group 4**

Mistrust of digital environments was frequently discussed, including fears about data misuse, hacking, and misinformation. Some participants recalled past negative experiences such as receiving fraudulent emails or being targeted by scams and expressed uncertainty about “*who’s getting hold of your information on the other side*” (**Participant 3, focus group 2**). This was summarised as “*an accumulation of things that have happened that alienate us against AI [artificial intelligence] and apps*” (**Participant 7, focus group 1**):

> *I don’t like putting my details on a system that other people can access. I don’t want to have messages from rogue people. I don’t want any communication like that*. **Participant 3, focus group 1**

This mistrust was compounded by concerns about the potential harms of digital technology more broadly, including the effects of excessive screen time (“*I also am very much aware of how long I spend on apps and on my phone all day, every day. I don’t want yet another thing that’s digital,*” **Participant 5, focus group 3**).

Despite these concerns, participants also recognised the potential value of digital tools. Some used general health apps to monitor diet or calorie intake, describing these as helpful for raising awareness and supporting behaviour change:

> *So for me, actually on this app was really useful for counting calories, because I was thinking like, okay, I will just put a bit of potatoes, a bit of meat, a bit of salad, and it’s like 300 calories, but then I’m just counting exactly and it’s 800. And now I just lost ten kilograms because I started to count actually calories*. **Participant 2, focus group 1**

Other participants, particularly those with type 1 diabetes, or type 2 diabetes with further complications (e.g., kidney disease), reported using sensors such as the Freestyle Libre to monitor their blood glucose levels which some described as “*life changing*” (**Participant 4, focus group 1**). However, even where such technologies were valued, some raised concerns about sensor accuracy, and becoming overly reliant on technology:

> *I do use a sensor, and I can’t tell you how fantastic they are. They do lie sometimes and they’re always about 20 minutes behind where I know I’m at. […] There is, I feel, a downside to the technology in terms of I now feel a little bit reliant on it. I don’t listen to myself as much*. **Participant 5, focus group 3**

Even when tools were perceived as beneficial, the practical demands of using them were sometimes described as “*overwhelming*” (**Participant 1, focus group 3**), particularly entering carbohydrate information which was experienced as burdensome and time-consuming, leading to frustration and disengagement. Participants also described confusion arising from the proliferation of apps with different functions and interfaces:

> **Participant 5**: *I think digital tools are good, but I think we’re just getting too many. […] If they could be integrated, so that there’s one sort of element, so that you can access one, I think that would be useful, cause it’s so disparate at the moment*.
>
> **Participant 7**: *You get you get 500 messages from every single one of them*. **Focus group 4**

This theme illustrates that digital engagement is not simply a matter of providing tools; it is shaped by a complex interplay of emotional labour, digital confidence, mistrust and the practical realities of diabetes self-management. Digital tools were often seen as potentially helpful but also potentially burdensome within an already demanding self-management context.

### Theme 2: The social conditions of digital self-management

Participants’ accounts highlighted that digital engagement was not solely an individual matter of personal capacity, but was embedded within the routines, relationships, and social contexts of everyday life. Digital tools were often experienced as constrained by structural limitations, including generic design, cultural mismatch, accessibility barriers, and financial constraints, all of which shaped how people used, interpreted, or avoided them.

Participants noted that many apps felt generic or “*made for the masses*” (**Participant 4, focus group 1**), and failed to account for co-morbidities, cultural dietary practices, or lived expertise developed through managing diabetes over time:

> *They might be suitable to some sisters, [but] they might not be suitable to me. I’m a heart patient. I have had open heart surgery. I’ve got high blood pressure. I have ulcers, so you know, there’s so many things I’m carrying along. So, if they [videos] say, tell me to eat something, I can’t do that*. **Participant 1, focus group 5**

The lack of cultural relevance was a particular concern. Some participants felt that digital tools did not reflect the diversity of people living with diabetes:

> *The cultural diversity that we have is not always reflected in the results and the information that is provided […] if you’re treating me like a five foot nothing White woman, and I’m six foot and I’m Black, it’s really different to the way that you would choose to treat me*. **Participant 3, focus group 1**

These concerns reinforced participants’ emphasis on their own lived expertise (“*we know our own bodies better than anybody else will do,*” **Participant 5, focus group 3**).

Accessibility challenges further limited digital engagement. Participants with dyslexia, limited English proficiency, or low health literacy described many digital tools difficult to navigate. Others lacked access to appropriate devices, struggled with small text, or faced financial pressures making digital support impractical or unusable:

> *Of course, if somebody was to show you how to do something on a laptop or a computer, that’s all very well, but if you then get home and you find, ‘Oh actually, I haven’t got a fast enough phone or a fast enough laptop,’ well, it’s a bit pointless really*. **Participant 1, focus group 1**

Financial costs created additional barriers, particularly for those on lower incomes and when devices needed to meet specific technical requirements, highlighting how digital health interventions risk reinforcing existing inequalities:

> **Participant 7:** *So you’ve got to have an up-to-date phone that will actually accept that app?*
>
> **Participant 4:** *Yeah, it’s got to have the NFC [Near Field Communication], like the same thing that you use for paying with the phone. So, it’s got to have the NFC and that’s pretty much all*.
>
> **Participant 7:** *It’s not cheap*.
>
> **Participant 4:** *Well, you can get a phone like this probably for like eighty quid, I would say*.
>
> **Participant 7:** *Yeah, but if you’re on benefits, you haven’t got eighty quid*. **Focus group 1**

These accessibility challenges reflected the social, economic, and environmental conditions shaping people’s opportunities, resources, and capabilities to access online and digital support.

Because of these barriers, many participants relied on family members, often adult children, to access test results, interpret information, or manage medication. This reliance reflected the extent to which digital tools often assume a level of confidence, literacy, and independence that not all users possess:

> *And the other thing is, you know, like we were saying that – we’re not very familiar with apps and everything, yeah? […] But one way that – you know, when we rely on the kids, right, kids are too busy with their own personal lives. They’ve got time, they will do it for you. If they haven’t got time, ‘Okay Mum, okay, I’ll do it in a minute*.’ **Participant 3, focus group 2**

Participants also valued hearing about the experiences of others, whether friends, family, or online communities. These accounts provided reassurance, emotional support, and a sense of shared understanding that digital tools alone could not offer:

> *The digital side of it. I don’t know, it quite sort of scares me because I like this sort of support, you know, talking to another diabetic. I don’t think you can get better than that. And I think sometimes, yeah, I can feel sort of quite isolated. And it, like, it’s not another person, it’s just an app. […] I think when these sorts of new digital stuff is coming out, it’s like, ‘oh, something else I’ve got to manage on my own.’ […] If you’re in a crisis, an app’s not going to save you*. **Participant 3, focus group 3**

Some participants were open to engaging with online peer support (e.g., via support forums, or online meetings), though anonymity and the lack of visible identity reduced trust for some. Even so, participants described the value of having access to others with lived experience:

> *…sort of a 24-hour thing where people maybe can get access to actually speak to other people if you’re really, you know, feeling that you can’t deal with it. […] It’s not easy and it’s hard when you haven’t got anyone to talk to about it, who’s going through the same thing*. **Participant 1, focus group 3**

Overall, digital self-management was described as a socially situated practice shaped by the wider conditions of people’s lives. The barriers participants identified, including limited English proficiency, low health literacy, lack of suitable devices, and financial pressures, reflected broader social determinants of health. Engagement with digital tools was therefore mediated by social networks, cultural context, and embodied knowledge. While digital tools were valued for monitoring and information, they were not seen as substitutes for the reassurance, interpretation, and shared understanding provided by family, peers, and lived experience.

### Theme 3: Healthcare structures shaping digital engagement

Participants’ engagement with digital tools was strongly shaped by their relationships with healthcare professionals (HCPs) and experiences of the wider healthcare system. Trust in digital technologies was closely tied to trust in the institutions and professionals who recommended and implemented them. Digital tools were welcomed when they enhanced access, communication, and continuity of care, but resisted when perceived as substitutes for person-centred, relational support.

Participants described significant variation in access to primary care depending on where they lived, with some reporting long waits and difficulties securing appointments for their diabetes management. Within this context, digital tools such as the NHS app were valued for enabling rapid access to test results and facilitating communication with HCPs. Participants recognised that digital tools could help relieve pressure on overstretched services and appreciated the convenience of accessing information when needed across multidisciplinary care pathways:

> **Participant 4**: *Yes, and the [cancer hospital] did have a specialist diabetes team […] your readings get transmitted to a team of your choosing. […] So, you can ring them up, talk about a problem. They just pull up your graphs and come up with a recommendation as to how to handle things*.
>
> **Participant 8**: *See that’s good use of technology*. **Focus group 4**

However, this appreciation was tempered by suspicion and concern. Some participants felt that digital tools were being used to push commercial or medical agendas, or to promote an “*arm’s-length approach*” (**Participant 5, focus group 4**) that prioritised efficiency over person-centred care:

> *I think you can be a bit suspicious when you think of what NHS trends might be in the next ten or twelve years, with more and more maybe American influence, and you wonder how accurate or impartial the information might be, or is it leaning towards supporting a particular product that they want you to use*. **Participant 6, focus group 1**

Others speculated that artificial intelligence (AI) might take over in-person consultations altogether:

> *Very soon you won’t be seeing doctor in person. It’ll be AI sitting on the other side. So, you’ll have to get used to getting information, whatever you need from the computer, because that’s the future*. **Participant 1, focus group 5**

Such concerns reflected a broader anxiety that digitalisation could erode the relational aspects of care that participants valued most. Across all accounts, participants emphasised that in-person interactions offered forms of reassurance, emotional connection, and non-verbal communication that digital tools could not replicate:

> **Participant 3:** *When somebody is talking, you know, even if it’s not in words, right, you know, your face impressions, you get all the message. So, with online, yes, yeah, they give you this and that, but you know when you’re – you don’t need to say in words, but, you know, your face is enough.*
>
> **Participant 1:** *You can better understand, that’s what she’s trying to say*. **Focus group 2**

Participants accordingly stressed the importance of retaining a choice between face-to-face or digital support, depending on their own needs and circumstances. A key barrier to digital uptake was the lack of support from HCPs in explaining how to use tools or why they might be beneficial. Simple signposting was seen as insufficient:

> *I’m a person that – if you give me a sheet and it says, ‘Do this, do that,’ it means nothing to me, but if you stand there and go, ‘If you do this and you do that and you do that,’ I will take it in. But if you give me a sheet and say, ‘Steps one to ten,’ by two, I’ve lost interest*. **Participant 7, focus group 1**

In the absence of such support participants felt uncertain, unsupported, and less inclined to engage with digital tools:

> *I think they’ve introduced technology, but they haven’t educated the people to use the tools that they’ve brought in […] I think it’s just been, ‘Here you go get on with it.’ I think that’s the wrong way to approach it*. **Participant 8, focus group 4**

Despite this, many participants were willing to learn, provided that digital tools were explained in ways that connected to their own needs and circumstances. Participants wanted clearer communication from HCPs about the personal relevance and potential benefit of specific tools so they could make informed choices about what support best suited their self-management:

> *But the NHS don’t tell you and the doctors don’t. They just assume you know what you’re looking for*. **Participant 1, focus group 1**

This theme demonstrates that digital engagement is mediated by trust in the healthcare system. Participants were more likely to embrace digital tools when they were perceived as enhancing communication, improving access, complementing existing relationships, and introduced with adequate support and explanation. In contrast they resisted digital approaches that appeared to replace relational care or reduce opportunities for meaningful interaction.

Taken together, these findings suggest that the acceptability of digital self-management is shaped by a dynamic interplay of personal capacity, social context, and trust in healthcare staff and services. Participants’ willingness to engage with digital tools depended not only on digital skills, but also on whether these interventions aligned with their lived realities, cultural contexts, and relationships with HCPs. Table 2 maps the data to constructs of the Theoretical Framework of Acceptability. Supplementary Material 4 provides further illustrative quotes for each theme.

**Table 2.**
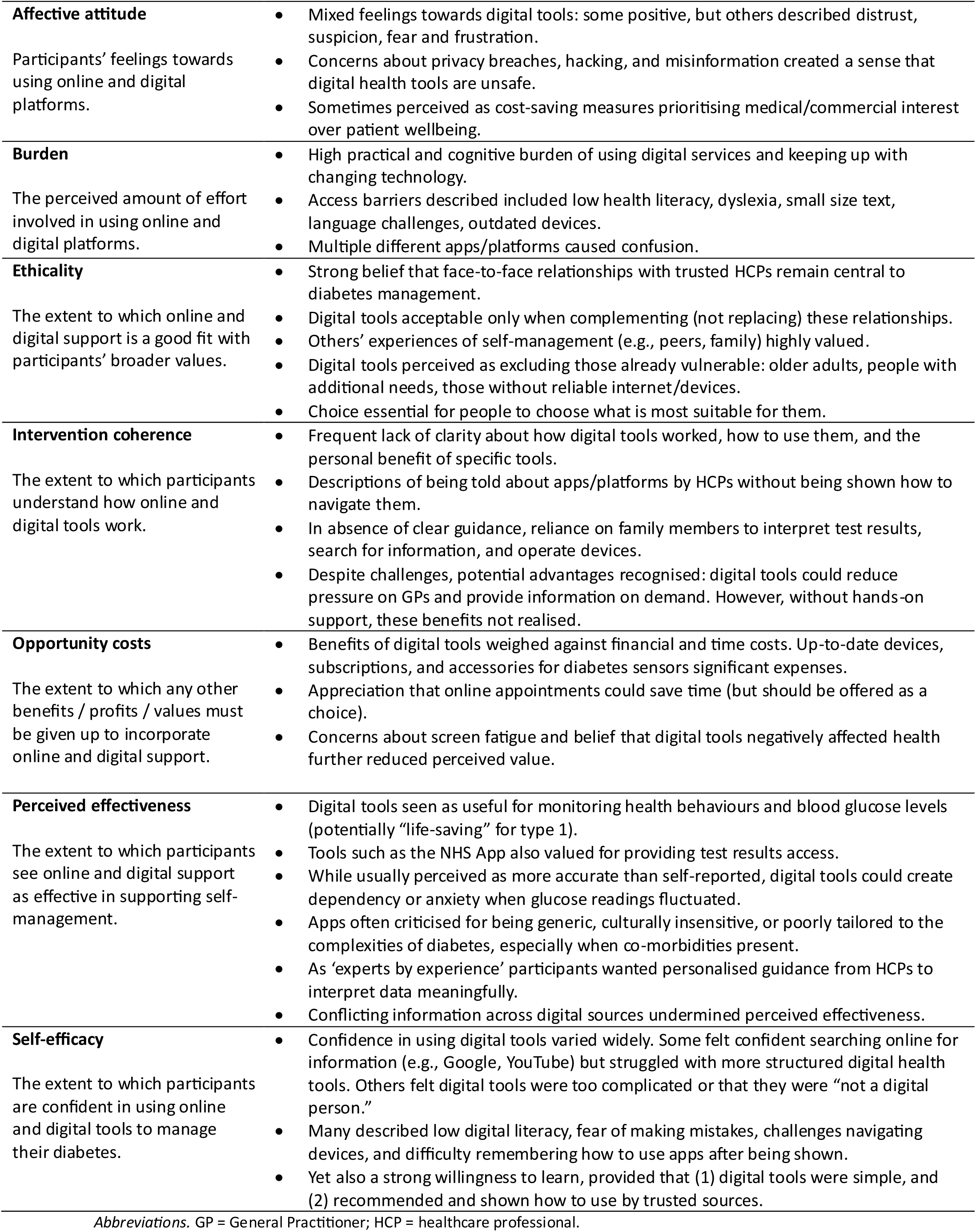
Data mapped to the Theoretical Framework of Acceptability constructs.

|  |  |
| --- | --- |
| **Affective attitude**<br>Participants' feelings towards using online and digital platforms. | Mixed feelings towards digital tools: some positive, but others described distrust, suspicion, fear and frustration.<br>Concerns about privacy breaches, hacking, and misinformation created a sense that digital health tools are unsafe.<br>Sometimes perceived as cost-saving measures prioritising medical/commercial interest over patient wellbeing. |
| **Burden**<br>The perceived amount of effort involved in using online and digital platforms. | High practical and cognitive burden of using digital services and keeping up with changing technology.<br>Access barriers described included low health literacy, dyslexia, small size text, language challenges, outdated devices.<br>Multiple different apps/platforms caused confusion. |
| **Ethicality**<br>The extent to which online and digital support is a good fit with participants' broader values. | Strong belief that face-to-face relationships with trusted HCPs remain central to diabetes management.<br>Digital tools acceptable only when complementing (not replacing) these relationships.<br>Others' experiences of self-management (e.g., peers, family) highly valued.<br>Digital tools perceived as excluding those already vulnerable: older adults, people with additional needs, those without reliable internet/devices.<br>Choice essential for people to choose what is most suitable for them. |
| **Intervention coherence**<br>The extent to which participants understand how online and digital tools work. | Frequent lack of clarity about how digital tools worked, how to use them, and the personal benefit of specific tools.<br>Descriptions of being told about apps/platforms by HCPs without being shown how to navigate them.<br>In absence of clear guidance, reliance on family members to interpret test results, search for information, and operate devices.<br>Despite challenges, potential advantages recognised: digital tools could reduce pressure on GPs and provide information on demand. However, without hands-on support, these benefits not realised. |
| **Opportunity costs**<br>The extent to which any other benefits / profits / values must be given up to incorporate online and digital support. | Benefits of digital tools weighed against financial and time costs. Up-to-date devices, subscriptions, and accessories for diabetes sensors significant expenses.<br>Appreciation that online appointments could save time (but should be offered as a choice).<br>Concerns about screen fatigue and belief that digital tools negatively affected health further reduced perceived value. |
| **Perceived effectiveness**<br>The extent to which participants see online and digital support as effective in supporting self-management. | Digital tools seen as useful for monitoring health behaviours and blood glucose levels (potentially "life-saving" for type 1).<br>Tools such as the NHS App also valued for providing test results access.<br>While usually perceived as more accurate than self-reported, digital tools could create dependency or anxiety when glucose readings fluctuated.<br>Apps often criticised for being generic, culturally insensitive, or poorly tailored to the complexities of diabetes, especially when co-morbidities present.<br>As 'experts by experience' participants wanted personalised guidance from HCPs to interpret data meaningfully.<br>Conflicting information across digital sources undermined perceived effectiveness. |
| **Self-efficacy**<br>The extent to which participants are confident in using online and digital tools to manage their diabetes. | Confidence in using digital tools varied widely. Some felt confident searching online for information (e.g., Google, YouTube) but struggled with more structured digital health tools. Others felt digital tools were too complicated or that they were "not a digital person."<br>Many described low digital literacy, fear of making mistakes, challenges navigating devices, and difficulty remembering how to use apps after being shown.<br>Yet also a strong willingness to learn, provided that (1) digital tools were simple, and (2) recommended and shown how to use by trusted sources. |

*Abbreviations.* GP = General Practitioner; HCP = healthcare professional.

## Discussion

### Summary of Findings

This study found that acceptability of online and digital support for diabetes self-management is shaped by interconnected personal, social, and healthcare system influences. Participants’ digital engagement was constrained by the emotional and practical demands of living with diabetes and by their own confidence in using digital tools. These individual challenges were compounded by social and structural barriers, including cultural mismatch, limited literacy, accessibility issues, and financial constraints. Engagement was further mediated by trust in healthcare staff and services: digital tools were welcomed when they complemented relational, person-centred care, but resisted when perceived as replacing it. While the Theoretical Framework of Acceptability (TFA) conceptualises acceptability in terms of individual perceptions (e.g., affective attitude, burden, coherence), our findings show how these perceptions are further shaped by structural barriers, social contexts and trust in healthcare services: acceptability was contingent on digital tools being embedded within a supportive, trustworthy, and person-centred healthcare system. Together, these findings highlight that engagement with digital self-management is not solely an individual choice but is shaped by social and healthcare system contexts.

### Comparisons to Previous Literature

Across the current literature, several qualitative and mixed-methods studies highlight consistent barriers to the uptake and sustained use of digital tools for diabetes self-management. A qualitative study involving adults from the United States with diabetes who owned smartphones but did not use digital health tools identified concerns about privacy and data security [38]. Another qualitative study involving people with type 2 diabetes living in Australia who identified as ‘app users’ or ‘non-app users’ found that barriers to use included a lack of endorsement or recommendations from a HCP [39]. Similarly, a mixed-methods study conducted in the Netherlands, which included both users and non-users of apps, found that participants wished to use apps collaboratively with their HCPs, but reported similar privacy and security concerns. Additional barriers included the time and cognitive effort required to track data, difficulties understanding app functions, limited professional support, and lack of financial coverage or reimbursement [40]. Interviews with people with diabetes from India who were already using online tools revealed that limited digital literacy, particularly among older adults, restricted independent use and created reliance on younger family members or friends to access online self-management information [41]. Our findings extend this evidence in a UK setting by showing how these barriers intersect with cultural relevance, family reliance, healthcare trust and concerns that digital tools may replace personalised face-to-face care.

Systematic reviews reinforce these findings, highlighting socioeconomic constraints, limited smartphone access, data costs, unstable internet, privacy concerns, and the burden of time-consuming technologies as key barriers to adoption and sustained engagement [13,42]. A recurring theme across systematic review evidence is the critical role of HCPs. Weak endorsement from clinicians and poor integration of digital tools into routine care consistently reduced motivation for people with diabetes to adopt or maintain use of digital support [13,43].

### Implications for Practice

Findings from this study highlight that people’s opportunities to engage with online and digital support to manage their diabetes are shaped by structural accessibility, cultural context and social support. Accessibility issues such as dyslexia, limited English proficiency, low health literacy, lack of appropriate devices, and financial pressures are not isolated individual challenges but reflect broader social determinants of health. These constraints are particularly relevant in diabetes because people from minoritised groups experience both higher prevalence of type 2 diabetes [44,45] and greater exposure to adverse social and structural determinants of health, including socioeconomic deprivation, discrimination, limited social support [46–48], and reduced access to high-quality diabetes care [49–51]. Digital health therefore risks widening inequalities if these determinants are ignored.

Digital diabetes support should therefore be culturally relevant, usable across literacy and language needs, and compatible with a range of devices. Digital options can improve access to information and support self-management, but only when they are introduced with appropriate explanation, demonstration and ongoing support. They should enhance, not replace, high-quality care. As the NHS in England continues to shift from analogue to digital services [15], maintaining face-to-face support remains essential so that people who cannot or choose not to use digital tools are not disadvantaged.

Uptake is likely to depend on how tools are introduced. HCPs should explain their relevance, demonstrate their use, and offer opportunities for follow-up, and will require sufficient time and training to do this. Rollout should be gradual, supported and tailored to individual needs, rather than adopting a one-size-fits-all approach. Working with representatives from older adults, younger people, and diverse communities can ensure that communication channels, language, and onboarding processes align with how people already use digital technologies. Community organisations, patient groups, faith settings and local leaders can act as trusted partners in promotion, onboarding and ongoing support, helping digital tools feel familiar, relevant and grounded in lived experience.

Financial and digital inequalities should be treated as core design and delivery considerations, particularly as self-management of diabetes is challenging for people experiencing socioeconomic deprivation due to financial barriers surrounding healthcare, and lack of access to devices and healthy food [52]. Digital interventions should not be standalone platforms but embedded within real-world systems that combine digital tools, face-to-face care and community-based support.

### Implications for Research

Future research should prioritise people most affected by digital exclusion to ensure that digital health innovations do not exacerbate inequalities [10]. This requires ongoing co-design with diverse communities to develop culturally relevant content, accessible delivery modes [53], and support that reflects how people already access and use healthcare [14]. Engagement should extend beyond those easiest to reach and include people who are typically under-represented in research. Partnering with community and religious organisations to co-design content, messaging, and support can help ensure that digital services reflect real experiences and enhance relevance for people living with diabetes.

### Strengths and Limitations

Whilst previous literature has examined the experiences of engaged users of digital DSMES programmes implemented in routine care (e.g., [23,24]), little attention has been paid to the perspectives of non-users. To our knowledge, this study provides one of the first in-depth qualitative explorations of why people with diabetes in the UK choose not to engage with online or digital support, such as MyWay Diabetes. Detailed descriptions of participants’ sociocultural context are provided to allow readers to judge the applicability of findings to their own settings. While the specific strategies identified may not be directly transferable to other minority ethnic communities or health conditions where different barriers may operate, the underlying process by which community-held knowledge is mobilised and sustained may offer a transferable framework for future work with other underserved populations. Further research is needed to establish the extent to which these findings resonate across other ethnic groups and long-term health conditions.

Close collaboration with PPIE contributors throughout enhanced the rigour and relevance of the research. Community leads helped develop study materials (e.g., study advert and topic guide), supported recruitment for three focus groups, and were present during those groups to provide translation and reassurance where required. Their involvement may have influenced who chose to participate. Nonetheless, they were trusted advocates who were committed to involving individuals who had not previously taken part in research. Their presence fostered trust and reassurance and contributed to a respectful and feasible methodological approach. This approach highlighted the importance of investing time in trust-building with community organisations and maintaining flexibility in research processes.

## Conclusions

Acceptability of digital diabetes self-management support is shaped by a complex interplay of personal capacity, social context, and healthcare system factors. Digital tools were valued when they enhanced understanding, supported self-management and complemented existing relationships with HCPs, but were resisted when they increased burden, lacked cultural relevance, or replaced personalised face-to-face care. Digital health interventions must therefore be equitable, usable, accessible, culturally relevant and embedded within trustworthy, person-centred care. As digital health becomes increasingly embedded in diabetes care, attention to these contextual factors will be essential to avoid widening inequalities and ensure that digital innovation delivers meaningful benefits for people living with diabetes.

## Declarations

## Acknowledgements

With particular thanks to the MyWay Diabetes Advisory Group (Dennis, Malcolm, Vivienne, Bonnie, G and Denis) who shaped all aspects of the research process, from the study design to data analysis and interpretation. We thank the staff of the community organisations for their assistance with recruitment and translation during focus groups, and the community members who participated for their invaluable contributions to this research.

## Ethics Approval Statement

This study was performed in accordance with the [Declaration of Helsinki](https://www.wma.net/policies-post/wma-declaration-of-helsinki/) and reviewed and approved by The University of Manchester Proportionate Research Ethics Committee (ref: 2025-23064-42006). Data presented in this manuscript is pseudonymised.

## Participant Consent Statement

Full consent was obtained from all participants prior to taking part in a focus group. As a ‘thank you’ for taking part in this research, people with diabetes who took part could opt to receive a £25 voucher for compensation.

## Data Availability Statement

A fully anonymised dataset (anonymised transcripts) is available in the open data repository Figshare at the University of Manchester Library: https://doi.org/10.48420/33288126.

The qualitative template analysis and previous versions of the template are available in the open data repository Figshare at the University of Manchester Library: https://doi.org/10.48420/33986137.

## Authors’ Contributions

MKR, DJW, DA, AB, HO-K, SGC, TW and JB secured funding for this project. JB, REH, AB, HO-K, MSB, TW, MKR and DJW contributed to the study design. REH led on submitting for ethical and regulatory approvals, data collection, data analysis, and writing the manuscript. CEFR and DMAA contributed to conducting the data collection. JB conducted the data collection, data analysis, and supervised the research conduct. DT, DA and REH led the Patient and Public Involvement for this work. All authors helped to prepare the manuscript and read and approved the final manuscript.

## List of Abbreviations

AI: Artificial Intelligence
DSMES: Diabetes Self-Management Education and Support
HCP: Healthcare professional
NHS: National Health Service
NICE: National Institute for Health and Care Excellence
REN: Research Engagement Network
TFA: Theoretical Framework of Acceptability
UK: United Kingdom

## List of Supplementary Material

Supplementary Material 1: Focus group recruitment strategy and procedures (.doc).

Supplementary Material 2: Focus group topic guide (.doc).

Supplementary Material 3: Patient and Public Involvement and Engagement (.doc)

Supplementary Material 4: Additional illustrative quotes corresponding to each theme (.doc)

