## Supplementary material for "Understanding the acceptability of digital diabetes self-management tools among non-users: A qualitative study of personal, social and healthcare contexts"

### Supplementary Material 1: Focus Group Recruitment Strategy and Procedures

The research team initially met with contacts from the Research Engagement Network (REN) in each of the three geographical areas to discuss recruitment strategies in their localities. The REN supports connecting researchers with Voluntary, Community, Faith and Social Enterprise organisations to better support diverse communities to get involved with research. Following this, we met with community leaders of various organisations and diabetes support groups, to establish whether they had members of their communities who were eligible to take part in this study. We aimed to focus on speaking with groups (1) whom diabetes disproportionately affects, (2) who are less likely to use online and digital services to manage their health, and (3) who are less likely to take part in research.

We developed relationships with various local community groups via (1) online meetings with the community leaders/trusted advocates, and (2) via engaging with in-person health events organised by community leads in their local community. The community leaders distributed information about the study, including the study advert and information sheet, to members of their groups who were eligible to participate, and acted as a 'mediator' between the research team and people in their communities living with diabetes.

We liaised with the community leaders about whether to conduct the focus groups online or in-person. Five out of the six focus groups took place in-person in participants' local community venues, organised by their community leaders. One focus group took place via Microsoft Teams. Focus groups were led by two female researchers who had training in qualitative methods. The aims of the study were described as wanting to understand participants' views and barriers to using digital and online support to help manage their diabetes. The community leaders were also present at three of the five in-person focus groups, who assisted with (1) translation where required, (2) helping participants to understand the consent form, and (3) providing reassurance and a friendly face for participants on the day. Participants provided full consent prior to the group discussion taking place.

### Supplementary Material 2: Focus Group Topic Guide

1. Please can you each briefly introduce yourself: what is your name, and what is your favourite food?
2. What do you think about the move towards using digital tools (e.g., apps) to manage health conditions?
    a. What do you think about using digital tools to manage your diabetes?
3. What are your experiences of using any digital services to help manage your diabetes?
    a. What do you like about them?
    b. What don't you like about them?
4. How easy / difficult is it to use digital support services for your diabetes self-management?
    a. What do you find difficult?
5. What do you think digital services for diabetes self-management are trying to achieve?
6. What gets in the way of you using digital support to self-manage your diabetes?
7. How do you think digital services would help with your knowledge of diabetes?
8. How do you think digital services would help you to manage your diabetes?
9. How confident are you in your ability to manage your diabetes?
10. How confident are you about managing your diabetes using digital services?
    a. What would help improve your confidence?
11. Is there anything else important that we haven't covered and you would like to talk about?

### Supplementary Material 3: Patient and Public Involvement and Engagement

*Reported using the GRIPP2 short form reporting checklist (Staniszewska et al., 2017).*

| Section and topic | Description |
|---|---|
| **1. Aim**<br>Report the aim of PPIE in the study | The aim of the PPIE group was to ensure that the entire research programme was informed and shaped by people with lived experience of diabetes. For this qualitative work, they helped to shape: the study design (topic guides, study adverts), methods of conducting the focus groups, interpretation of qualitative findings, and the recommendations arising from this work. |
| **2. Methods**<br>Provide a clear description of the methods used for PPIE in the study | Members of the PPIE group included n=6 people living with type 2 diabetes, across the three study sites and including a range of MyWay users, non-users and past users:<br>• n=3 female, n=3 male<br>• n=1 African Caribbean, n=1 Asian British Chinese, n=1 Asian Indian, n=1 Asian Pakistani, n=2 White British<br>The PPIE group met with the research team via two-hour videoconferencing meetings every 2-3 months throughout the duration of the research (18 months).<br>This work was designed with a Public Co-Chair who met the PPIE and research lead in fortnightly project meetings, taking part in quarterly project management board meetings.<br><br><u>Shaping the focus group questions</u><br>• Contributors provided feedback on the wording and order of the focus group questions. Topic guide was circulated via email in advance of a two-hour online meeting.<br>• During the meeting, the group reflected on:<br> o Are these questions clear and open enough for people with different experiences?<br> o Could we change or reword any questions to feel more clear or relevant to everyone?<br> o Do they invite people to reflect on what matters most to them?<br><br><u>Shaping the study advert for focus group recruitment</u><br>Group were asked to provide their feedback on the draft study advert via email. Group were asked:<br>• Is it clear what the study is about?<br>• Is anything unclear or potentially off-putting?<br>• How could the advert be made clearer, more attractive, or more accessible – particularly for people who might not usually take part?<br>• Where might we consider promoting the advert to reach a wide range of people?<br><br><u>Shaping the conduct of the focus group discussions</u><br>• Discussion on how to make focus groups feel inclusive and encourage people to share their views during the focus groups.<br>• During the same two-hour online meeting as shaping the topic guide questions, the group discussed:<br> o What helps you feel comfortable sharing in a group of new people?<br> o What might help others feel safe and open in that situation?<br><br><u>Interpretation of the qualitative focus group findings (with MyWay users)</u> |

| | |
|---|---|
| | Contributors were sent one pseudonymised transcript (via either email or post) and were given approx. 6 weeks to read and comment on the transcript.<br>They were sent a document called 'How to read through workshop transcripts' which explained what they were being asked to do, offered guiding questions to help them reflect on the transcript, and provided some tips on how to make reading the transcript easier.<br>When reading the transcripts, contributors were prompted to reflect on the following guiding questions and send back their reflections prior to a group meeting:<br>1. What were your first impressions – what stood out most and why?<br>2. What were important and/or common benefits, challenges, or ideas raised by participants?<br>3. What do you think are key points for people to take from this transcript?<br>4. Is there anything else you would like to let us know?<br>During a subsequent two-hour online meeting, the PPIE group together discussed:<br>1. What stands out to you as the most important, surprising, or meaningful insights from the transcripts and themes?<br>2. Are there experiences, concerns, or voices that feel missing, underplayed, or less visible in these themes?<br>3. How do these reflect real experiences of people living with diabetes, and what might the practical implications be for people's daily lives?<br><br><u>Shaping the recommendations arising from this work</u><br>• During a two-hour group meeting, researcher presented draft recommendations for the MyWay team specifically, based on research findings. Group discussed the following:<br>1. What do you think are the key things [researcher] has captured and is that reflected?<br>2. Are there important themes or perspectives missing or underrepresented?<br>3. What comments do you have on these as a whole based on our work up to now?<br>• The group then split into two breakout groups and discussed the above questions in more detail. |
| **3. Study results**<br>Outcomes – report the results of PPIE in the study, including both positive and negative outcomes | The PPIE group shaped this research in the following ways:<br><br><u>Shaping the focus group questions</u><br>The group commented on the wording of the topic guide questions. Suggestions included:<br>• Adding in an ice breaker question asking about favourite foods which would not be time consuming but put participants at ease.<br>• Adding in a prompt question about confidence in using technology/online platforms.<br><br><u>Shaping the study advert for focus group recruitment</u><br>The group suggested changes to the study advert wording, including:<br>• Changing wording of 'digital services' which people won't understand.<br>• Changing the wording of "joining a focus group" to "joining a group discussion."<br>The group suggested clarifications in the advert, including:<br>• Make clearer why people might want to take part, e.g., "You can make a difference by…" |

- The target audience (i.e. interested in speaking with people living with both type 1 and type 2 diabetes).

The group provided suggestions on the images used in the advert:

- As well as picture of smartphone, include a picture of a laptop, tablet, desktop etc.
- Increasing diversity of people shown in images, e.g., include woman with headscarf, have representation of older people.

The group suggested ways to engage and recruit:

- Suggestion for the researcher to be present at community group meetings/information days to give a more personal touch.
- Some contributors had contacts of relevant diabetes support groups in their geographical areas who they linked up with the research team to help with recruitment.

<u>Shaping the conduct of the focus group discussions</u>

The group provided the following suggestions to make the focus groups as inclusive and open as possible:

- For the in-person focus groups:
    - Suggestion to share the topic guide with the community leaders prior to the focus groups so that they could explain to potential participants what questions would be asked and help with any uncertainties that participants may experience beforehand.
- For the online focus group:
    - Suggestion for researcher to send short video pre-focus group to set expectations and help personalise it for participants, helping them to feel relaxed and save time during the focus group.
    - Suggestion to offer participants a practice session using Microsoft Teams before the scheduled focus group for anyone not feeling confident.
    - Suggestion to ask participants to mute their microphone when others are speaking, as neuro-diverse participants can find background noise difficult and distracting.
    - Discussed that the chat function in online meetings can be distracting; suggestion to make clear to participants that they do not have to follow what is said in the chat (this is the researcher's job).

<u>Interpretation of the qualitative focus group findings</u>

The group shared their reflections on the transcript analysis activity, and discussed:

- Users not being shown how to use online tools.
- How people are supported to engage with online and digital services.
- Addressing inequalities using a more targeted approach.
- How this links up to real life services and communities and interest group (infrastructure).
- Digital tools are positive but not a replacement for face-to-face services.

<u>Shaping the recommendations arising from this work</u>

The group discussed recommendations arising from this work:

- Results give the MyWay Digital Health team a chance for reflection.
- Accurate connection to real time NHS data is essential for take-up, trust and engagement
- A clearer link to the NHS in promotion and onboarding supports sign-ups as the NHS is a trusted name
- MyWay need to refine their product in a more tailored way.
- MyWay need to find a way to make their platform more personalised.

| | |
|---|---|
| | • MyWay need to involve the people they want to reach in doing that.<br>• Who are the people that these platforms need to target and how to do that effectively rather than trying to appeal to everyone.<br>• Addressing inequalities using a more targeted approach, e.g., working closely with local communities and organisations.<br>• It needs to be made clearer to potential users what the benefits are of using these online and digital tools.<br>• Digital exclusion is an essential long term goal to be addressed |
| **4. Discussion and conclusions**<br>Outcomes – comment on the extent to which PPIE influenced the study overall. Describe positive and negative effects | Outcomes of PPIE input throughout this study include:<br><br>• The focus group topic guide was amended and shortened based on feedback.<br>• The ice-breaker question in the topic guide was based on suggestions from the PPIE group, which provided a shared starting point for participants.<br>• Changes were made to the wording of the study advert based on the group's feedback to enhance clarity.<br>• Images used in the study advert were diversified and different images were tailored for different community groups.<br>• For the online focus group, researcher provided participants with an introductory video, including how to use Microsoft Teams, ground rules and list of topics that will be discussed during the focus group – this helped to keep focus group to time on the day, and we received positive feedback from research participants that the introductory video aided their participation.<br>• Focus group 'ground rules' were shaped based on PPIE feedback.<br>• Participants were offered a practice session to use the technology before the scheduled online focus group.<br>• Discussions with the PPIE group have helped to shape the findings and ensure they also reflect perspectives of people with lived experience of diabetes.<br>• In addition to reflecting the patient voice within our academic papers, the key PPIE group priorities and recommendations arising from this research will be explicitly listed within the NIHR final report in detail. This report includes considerations around:<br> ○ Offline support to support people capable but not confident with technology to use the app.<br> ○ Greater consistency in national approach alongside place-based approaches tailored to their key publics and community assets.<br>• A member of the research team fed back the project findings and recommendations to the MyWay team; those recommendations were shaped following discussions and feedback from the PPIE group.<br>• The PPIE group will have a feedback session directly with the MyWay Team in late 2027 as a chance for the MyWay Team to respond to comments and recommendations. |
| **5. Reflections/ critical perspective**<br>Comment critically on the study, reflecting on the things that went well and those that did not, so others can learn from this experience | PPIE has been vital throughout the life of the wider research project. The regular dialogue and positive rapport between the research team and PPIE group ensured that public contributors were able to input on the study design, methods, findings and wider implications of this work, which provided a unique and broader lived experience perspective.<br><br>A patient and public involvement and engagement manager from Vocal (https://wearevocal.org/) led the PPIE group, including recruiting PPIE members, communications via email, circulating relevant research materials and meeting agendas, developing and facilitating the online meetings (with a |

member of the research team present) and regular meetings with a member of the research team and public co-applicant every two weeks to plan the PPIE activity throughout the 18-month study duration. Having a dedicated person to coordinate the PPIE activities meant that public involvement was at the forefront of this work, ensuring that each ‘ask’ of the PPIE group was valuable to the research team, and the group had a key contact to raise any issues/queries/feedback throughout the project.

It was through these regular meetings that the decision was made to run an informal ‘book club’ transcript exercise with the PPIE group, proving an opportunity to develop their own interpretations off the data, rather than solely commenting on pre-formulated research recommendations. Group members reported that they valued this approach, describing increased feelings of empowerment and ownership over the research. The exercise also contributed to the development of patient-led recommendations, broadening the potential impact of findings. However, the exercise required varying levels of time and effort from different contributors, with some spending considerably longer on the task than others that the team only discovered later. This highlighted challenges in recognising and reimbursing time equitably and ensuring contributors feel able to ask for support during tasks.

PPIE contribution for a research evaluation of an NHS service already implemented had unique challenges. For example, it was not within the remit of the research team to directly make changes to the MyWay programme itself. This required careful management of expectations during onboarding, with a clear explanation of the group's role and remit. To support this, an introductory information pack was developed and shared with contributors, alongside individual introductory meetings. The first group meeting was dedicated to onboarding and establishing the purpose of the PPIE group.

The PPIE group were able to have impact on the service through shaping the research and their own recommendations, as well as direct contact with the service provider, who could provide feedback on the actions they would take as a result of this research. This was vital so that the PPIE group were informed about the difference that their contributions have made to this work.

### Supplementary Material 4: Additional illustrative quotes corresponding to each theme

**Table S1. Additional illustrative quotes corresponding to each theme.**

| **Theme 1: Diabetes self-management and navigating digital environments** |
|---|
| **Navigating diabetes management** |
| **P1:** No, no, no, it's not me physically taking them [tablets]. It's not the physically taking them. It's what it's doing to your body is what worries me. Having been somebody who, up until six months ago, was taking absolutely no medication whatsoever, I don't want to be taking all this medication I don't want to be taking – again, like you were saying, they say, "Oh, get onto statins." I said, "No, I don't want to be on statins." You know, so I'm on some blood pressure tablets, but I want to get off those as well. I want to work out how do I get off them [laughter], you know.<br>**P7:** Get off them blood pressure tablets [inaudible 0:57:10] [laughter], that'll cure your stress levels straight away.<br>**P3:** I don't know that the app will offer that information either. **Focus group 1**<br><br>**P3:** So, everyone is different, you know. But at the end of the day, right, I'll say this, even myself as well – I'm not blaming anyone, I'm sorry. But, you know, if you've got willpower, you can do anything. You have to have strong willpower, right? For example, right, I'm going to live for myself, I'm going to look after myself, I'm going to do healthy eating and all that, if you've got, you know, strong willpower, I think you can beat anything. But if you haven't got willpower, no matter – the doctor, the consultant, whatever, right, they can do whatever they want to do, you know, if you can't control yourself, right, that's it.<br>**F1:** So, how confident do we feel then around this table – how confident do you feel in managing your diabetes?<br>**P3:** I'm poor, I'm very poor. I can't control my diabetes. **Focus group 2**<br><br>**P3:** I think people used to be ashamed of having diabetes, medical conditions there is nothing to be ashamed of.<br>**P1:** Yeah, exactly. It's happened to you. It's happened to so many people out there. […]<br>**P1:** I think it's we Black that we feel ashamed of something that's happened to us. And by that, we hurt ourselves.<br>**P2:** That's true.<br>**P1:** Yeah, because if you hide this, you hide this, you hide this until it gets worse. **Focus group 6**<br><br>One thing is like if I wake up early morning, have my breakfast, do whatever I need to do. But during like about 12 or 1 o'clock, my legs go very weak. I cannot even, I couldn't fast these months, what we do in Ramadan. I couldn't fast because during about 1 o'clock in the midday, my legs go weak, my sugar level go very down. And then as soon as I eat something, then I feel a bit better. And this, it started happening to me, this is two years ago. Yeah. So, before I was fine, I used to keep all my fasts, everything, but now. **Participant 4, focus group 5**<br><br>**P1:** Yeah, maybe a list would be good. Someone like you, [P7], having a list of, "This is the foods that are reasonably okay," but not too many, just sort of a good list, "And these are the ones, oh, you need to watch the portion size on that," you know, that kind of thing.<br>**P7:** The thing is, on my budget, I tend to eat more stuff like stews and pasta, things like that, and I know some of them can't be good for you. I mean, I have cut down a lot on potatoes, because apparently they can be quite detrimental, but I do love potatoes [laughter]. Don't get me wrong, if someone said to me, "Do you want a Maccy D's chips or something?" Hmm [laughter], you know, that's the type of thing – but the thing is, it's cheaper to go to Maccy D's and buy a bag of their chips than it is to go to an organic place and buy their food. **Focus group 1** |

**P1:** I think you need to know about how much sugar it's got, how much calories it's got, because once you eat that sugar, you need to burn it. If you don't burn it, it stays in your blood. And that's what rises your, you know, levels of your sugar, because-
**P6:** Naan is not good for diabetic person.
**P1:** Roti is no good, but my doctor, you know chapati? Chapati, my doctor, the specialist in the hospital, he told me that, “Don't eat chapati.” I didn't eat chapati for six months.
**F1:** Oh wow, chapati is good.
**P1:** And that brought my sugar all the way down.
**P6:** Yeah, you just stop always.
**P1:** Yeah, chapati. I mean, I said to him, “I want chapati. I can't give up chapati.” He said, “There's millions of people out there don't eat chapati, they're not dead.” That's what he said to me, “They're not dead, you're not going to die.”
**P6:** White bread don't go, brown is okay. And then brown and chapati flour roti, as well. We only eat only this size [shows portion in hands].
**P3:** You think is chapati is not good for diabetes?
**P1:** Yes, because no, no, no, because chapati has high carbohydrates in it. It takes time for your body to digest them. So, you take, if you're taking carbohydrates, take something which digest quickly. And doctor says, this is Asian doctor, [name], and I've done it. And it does take you sugar less down. What you do is you take, go and have a bread with zero carbohydrates, zero sugar, and also pita bread. You could have pita bread, which is, yeah, zero. **Focus group 5**

**P1:** There was a time when I used to put sugar, obviously, like one, one and a half in tea, but I stopped all that. I even used to have tablets, I’ve stopped everything. I’ve got loads of sugar at home but I do not use it in the tea.
**P6:** What about sweetener?
**P1:** No sweetener, I stopped all that. I used to have it a year ago but I’ve stopped it, because I’ve had diabetes for a couple of years. But obviously, like I said, it’s like willpower, isn’t it? I stopped adding it to milk as well. You can do it. I’ve cut sugar. Like I said, I used to have one and a half sugar, or one maybe. I do put honey now and again, ‘cos I get a sore throat sometimes, so I use honey sometimes, but not every day.
**P6:** What about those sweetened regal teas and Chai?
**P1:** No, nothing, I stopped everything. Not good for my blood pressure as well, so it’ll go zooming, so it’s one thing or the other. But like I said, sugar, I’ve cut it. **Focus group 2**

**P1:** Yeah, like with our age, we need to be careful.
**P6:** Yeah, age is going by, you know, more healthy food and stuff. But it’s like a mixture really, because you can’t really keep on eating healthy. You have to eat –
**P3:** Other stuff [laughter].
**P4:** You don’t have to eat junk though [laughter].
**P1:** As well as we’re Asian, obviously, we eat a lot of spicy food.
**P6:** Curry dishes and stuff.
**P1:** I’ve cut down now. I used to do a lot of fried stuff, kebabs and that, and mostly – a lot of things I’ve cut down for many years now.
**P6:** Spices and stuff.

| |
|---|
| **P1:** Spices, I had always when I was young, when I was like before thirty, forty years. So, that's all that we had at a young age, now it's affecting us. Now I've stopped all that, you have to, otherwise I'd be bedridden, I would be.<br>**P6:** Yeah, we have to watch what we cook and the amounts we put in the cooking.<br>**P1:** The chillies as well, you know, red chillies, they say they're not good, so I've cut them down to one now. I used to put two in at a time. I've cut down to one now, so that's a bit better for me now. **Focus group 2**<br><br>My doctor says, you have to with the diabetes, you have to walk certain, like, you know, you have to make sure that you're walking one hour per day or two hours, whatever you can afford, you know, do it. But with the walking, I my, personally myself is very good because that gets your diabetes down, plus you're drinking a lot of water and that makes better. And when you're sitting at home, you're always craving something. You want to feel this, you want to feel, you see, you open your fridge, you see something, you started eating now. And if you spend your time busy doing something else, then you don't crave for anything like, and that way, it's helping yourself as well. **Participant 4, focus group 4**<br><br>Yeah, I've had it [type 1 diabetes] sort of like what, 5 and a half years now, and never having a problem with food at all, not having to really, being really lucky, not having to watch what I eat, [and then] all of a sudden it was like my life's just changed overnight. Yeah, so I still struggle with my sugar levels, I try and manage it the best I can, and I can do because I've got the sensor. But again, like Participant 5 just said, it's 20 minutes behind what the, like, your real sugar level is, and in 20 minutes, if you're having a hypo, that's a hell of a long time. **Participant 3, focus group 3**<br><br>Yeah, it was a lot of information to take on to start with, and I think, you know, you sort of panic to start with, and you think this is – you know [laughter], "you can't have this", and then you calm down a bit and think okay this is fine. I don't know. I did a lot of research, looking up stuff online. I mean, I just googled it. **Participant 1, focus group 1**<br><br>**P3:** Websites, you know. But sometimes, you know, the websites and the information can be different as well.<br>**F1:** Yeah, so tell me more about that.<br>**P3:** What do you call it, like for example if you type in Google, right, and say, you know, "What should be the normal rate for the sugar in diabetes," and all that, that will give you some different information. So, an expert, like a diabetic nurse or anything, they will give you something else, you know, different information, yeah, and then you get confused, "Who do I listen to? Do I follow the internet or do I listen to the diabetes nurse?" Or sometimes – you know, like all us ladies here, everybody experiences different ways with their diabetes and everything, yeah. So for example, if Participant 5's saying that, "Look, I do everything but my sugar level is level." Right, I said, "I'm doing everything, my sugar level is really high." But the thing is, what I'm trying to say is that everyone is different. But sometimes, you know, if somebody says, "Look, I've tried this and my sugar level has controlled," you do try it, you know. There's no harm in trying it, you know, with your limits, right? You can be either okay or you can get worse. So, like I said, at the end of the day, right, just try to control yourself if you can, you know, whatever food… **Focus group 2** |
| **'Not a digital person'** |
| I'm failing that because I don't have much knowledge about internet. I can read a message, but going through apps and doing things like that, I'm no good. **Participant 3, focus group 2** |

But the thing is because I never took IT when we were in school and everything, so I mean, with apps and websites – you know, that's something new, you know. That is for the youngsters, not us lot, you know. I mean, they say that you're never too old for anything, but I conclude, I don't think I can pick that up that quick. It's hard, very hard. It's very hard. **Participant 3, focus group 2**

**F1:** So, what is it that you would find hard if like you spoke to a GP and he said, "Go online and look at how to manage your diabetes?"
**P4:** I'd go and see my daughter, but for me, I can't.
**F1:** What is it that you would find difficult?
**P4:** To search myself, anything like that.
**F1:** Is it knowing what to search for or how to search?
**P4:** I'm going to wrong app and – because I've not got much knowledge – when I was in school, we had no IT, no nothing. **Focus group 2**

But like Participant 2 said, you do get used to what's in the food that you're eating. So, then you can administer your insulin according to what you're eating and, you know, sort of stuff to avoid if that's what causes your sugar levels to spike. So yeah, I'm not really a digital person. **Participant 3, focus group 3**

Yeah, that is, and I think what is going to affect some of people using the digital is confidence of using, like me! Confidence is, you know, if you are not a computer literate, it's going to be difficult. […] And it's going to be difficult like still say if you know when you get old you tend to forget things. **Participant 1, focus group 6**

**P6:** That's what's affecting your health as well, your eyes and –
**P1:** You're on it too much, on your phone too much.
**P6:** It's not good for your health.
**P1:** No, it's not good. **Focus group 2**

**P6:** At least you've got email. I've got no email, no Instagram, no Facebook, nothing.
**F1:** Oh, so you don't use email or anything.
**P6:** No, I don't.
**F1:** What's your reason for that?
**P6:** I'm not into all that. I use my son's email. He got me on and I go, "I'm not into all that." I wouldn't want to. I use my son's email. I don't like going on the phone.
**F1:** Is that just a personal preference or is it specific worries that you might have?
**P6:** No, just, like I said, you know, like your physical health, you know. You get headaches looking at the screen too much. Yeah, it's too much, too many things going on, and especially nighttime, a lot of people are on the phone at night, and it's meant to be bad, especially at nighttime. It's affecting your health. **Focus group 2**

Yeah, I also am very much aware of how long I... spend on apps and on my phone all day, every day. I don't want yet another thing that's digital. You know, I feel we have so much crowded lives already. Yes, another app. It's kind of almost in a way nice to have a break from it. **Participant 5, focus group 3**

I've got an Android phone which is very good, but I'm very – listen, I can hardly do anything. I try to get my son to help me, and he confuses me more, but I am very, *very* nervous at technology. If it's to do with diabetes, I'm really scared, I'll be honest with you. I've got no confidence with it. **Participant 3, focus group 4**

| |
|---|
| Because what is sometimes, you know, when you're using the computers or anything like that. What if you by mistake you click something else, but you're not supposed to and you don't know how to get it out. Sometimes we don't know how to use the computers and it's too late for us to learn. But when we do learn, it's the forgetfulness, you know, sometimes you forget as well. So, it's hard. **Participant 4, focus group 5** |
| **Distrust** |
| **P7:** It's all an accumulation of things that have happened that alienate us against AI and apps.<br>**P3:** I would agree. I would say knowing that what you're watching is AI – sometimes you're not sure what you're watching. Is that Oprah Winfrey or is it like some AI image, telling you that you can use those two injections that the staff are using at the moment? I can't remember what they're called, so it's very effective [laughter]. But the weight loss injections and the methods by which information has been fed out to us means that I am really reluctant to trust – and as you're saying, could it be used, could the results of this be used, "We now have 10,000 people using this app, and of those 10,000 people, there's those on Metformin, there's those on this, we can influence the market, because we can offer cheap medication that will deal with these 5,000 people." I don't want to be part of that. **Focus group 1**<br><br>I do use other apps that require very little information. So, I don't like putting my details on a system that other people can access. I don't want to have messages from rogue people. I don't want any communication like that. **Participant 3, focus group 1**<br><br>**P3:** I'd want to feel protected too, so that my information isn't going to go off –<br>**P7:** Oh yeah [inaudible 0:32:25] and loads of these want so much information, and then you get, "Oh, we do apologise, but we're having to send out a letter saying that, you know, we've been hacked and all your information has gone elsewhere." And the next thing you know is you're getting ten million adverts for items that you don't even want to know. **Focus group 1**<br><br>So, what I'm trying to say is that, right, if it's, you know, a simple way, you know, instead of relying on the app and all that, if it's on paperwork, a letter or face-to-face and all that, I think that will be better for us to understand, you know? Because I know everything's on a website or apps and all that, you know, it could be true, it could be false, you know, anything. But the thing is that if you don't know yourself how to do it or log in or anything, right – like Participant 4 just said that, somebody tried to hack her account, you know. So, us lot, we think that, okay, we put our details in and everything, yeah, you know, we done it okay, but you don't know who's getting hold of your information on the other side, you see. So, you know, this is how it is, you see. Because we think that, okay, yeah, what's your name, your address, your date of birth, and blah-blah-blah, you know, and we just – because the data is for ourselves, we're typing it, but we don't know, what do you call it, who's on the other side, you know. It could be anything. It could be criminals, for example, it could be fraud, and by the time you realise, all your bloody money's gone. **Participant 3, focus group 2**<br><br>But yeah, I am concerned about that [data privacy], you know, because just because of, you know, where I've worked and things like that, you know, I know that a lot of information is, you know, stored offshore and things like that and anybody can have access to it, you know, and okay, they tell us now, you know, this and that's going to be shared. And even with apps, if you actually go into any of your apps on your phone, you'll see that you've actually, without even knowing it, agreed to quite a few things and, you know, maybe things have been installed on your phone and you're not aware of it. So, I do worry a little bit about that. But at the same time, what can you do? It's like you've got these digital technologies and you kind of have to give those permissions and some of them, there's agreements where they can take that information without you even being aware of it. So yeah, it's a difficult one, but it's something that I worry about, you know. **Participant 1, focus group 3** |

| |
|---|
| I'm still really mistrustful of AI. And I, again, I go back to if you talk with somebody who's actually having the same experiences or, you know, the same condition, that just, you know, supersedes anything by AI for me. […] So, I don't know what could be done to make me trust it more because I kind of think, you know, the technology is great, but it's not good enough yet for me to really trust it. **Participant 5, focus group 3** |
| **Apps are useful for monitoring** |
| So for me, actually on this app was really useful for counting calories, because I was thinking like, okay, I will just put a bit of potatoes, a bit of meat, a bit of salad, and it's like 300 calories, but then I'm just counting exactly and it's 800. And now I just lost ten kilograms because I started to count actually calories. So that is helping me, because I didn't realise that actually that will be that much calories in the plate, yeah. So, for me, on this app, I just get like a one year, so it's really useful for counting calories, yeah. […] But for keeping a calorie count, keeping your body to move – like you get in the app, "Okay, stand up now like ten [inaudible 0:14:38]," or whatever, yeah, for me, it's useful. **Participant 2, focus group 1**<br><br>So, I'll use like maybe Huawei, is it Huawei or Samsung, something like that, literally just to sort of try and log the carbs, but then by sort of lunchtime, I've just lost the plot, you know, I can't. So, but I have found them useful because it's a good reminder. It keeps, you know, let's say the Huawei one then, because I walk a little bit. So that's helpful because it's got places to record stuff. So, I just kind of stick to that. So, it's not even specifically about diabetes, but the information that I input there, it does help. So I think even if you've got one, I think it can be helpful if they can, you know, tweak it a bit more. **Participant 1, focus group 3**<br><br>Okay, yeah, so mine is managed primarily by the Freestyle Libre sensor that I've got on my arm. Yeah, and that's sort of connected to my phone. So yeah, I can sort of set alarms and stuff, and it can give me warnings before my sugar level goes too high or too low. And I also use the Carbs and Cals app to, yeah, to calculate what sugars I'm eating and not eating, and what's got a lot of carbs in and stuff like that, so yeah. **Participant 3, focus group 3**<br><br>**P4:** I mean, there is a lot of useful apps. 'Cos most of you don't use LibreLink [app].<br>**P1:** No, no, I think you're the only one.<br>**P4:** For me, that was life changing, honestly, it was a life changer. It changed my control of diabetes totally. **Focus group 1**<br><br>And the technology that's developed over the past few years, I think has really helped me. I don't use a pump, but I do use a sensor, and I can't tell you how fantastic they are. They do lie sometimes and they're always about 20 minutes behind where I know I'm at. But I do feel that the technology that's coming on is great. There is, I feel, a downside to the technology in terms of I now feel a little bit reliant on it. I don't listen to myself as much. For example, I changed sensor yesterday and I gave myself 12 hours just to have a bit of a rest of having this bloody thing on my arm, you know. And I doubt myself because I'm so reliant on the technology. And I think that potentially is a little bit bad for me, because we know ourselves, we know our bodies better than anybody else will do. And you have to listen to yourself sometimes. So, technology has both pros and cons for me. Um, yeah. **Participant 5, focus group 3** |
| **Multiple apps cause confusion/overwhelm** |
| **P5**: I think um digital tools are good, but I think we're just getting too many. […] If they could be integrated, so that there's one sort of element, so that you can access one, I think that would be useful, cause it's so disparate at the moment.<br>**P7**: You get you get 500 messages from every single one of them. **Focus group 4** |

I was just going to say, like Participant 2, I've downloaded quite a few different apps, even Google ones. And yeah, the same thing, you sort of sign up, you know, start using them, and then you either forget or there's no prompt. This is what I'm finding, as well, for you to sort of go back and say, you know, oh, let me enter this. And yeah, it does become, I think, frustrating then having to put all that information in, you know, so I have, I took a few of them off the phone, but I find that I don't end up using them either. I don't know what it is really. **Participant 1, focus group 3**

**P4:** Then the other thing of that is finding time for this as well, yeah? 'Cos especially when you work, if you've got to spend like, you know, time in the morning to look at the apps or this or that, try to work out what you're going to eat – how you're going to eat or this, or other information – because we focus on diets and things –
**P7:** Then you've got to get up another half hour to an hour earlier. **Focus group 1**

**Theme 2: The social conditions of digital self-management**

**Perceived genericness of digital tools**

No, I don't agree with putting everything on an app. Certain things, yes, but medically, I don't think it would work properly, because it's too… Just a broad question mark. If you don't answer any of them questions, it doesn't affect you. Like I've had plenty of [inaudible 0:04:55] when I've actually phoned up or tried to make an appointment, and it says, "What's wrong?" You know, there's nothing on there that actually describes what's wrong with me. And then it says at the bottom, "Fill in what is…" And I'm not any good at writing or anything like that, so I'd rather be able to speak face-to-face or even over the phone with someone, explain what's wrong with me, and they can say, "Well, come in here," or, "Get to the hospital." Whereas an app will go, "Well, you don't reach our criteria, so…" **Participant 7, focus group 1**

Well, for me it's – I think it's a bit different because I'm type 1 and much longer, so I use my experience a lot, and I look in some apps, let's say especially the diet ones, counting the calories and everything, but then personally I think it's – apps are made for masses, for loads of people. They're not personal at all. So if you think about – every person will be totally different. Especially looking myself, there is much more stuff than just diabetes now. I've got a feet problem, some nerve damage and things like that, so I'm taking other medications that actually – I know they affect me. Doctors said they shouldn't. I know they do because I know how my sugar goes, and it's not going to come from nowhere, it comes from somewhere. So, using anything like that – some stuff is useful, but then you have to take on board what else happening with you, because it's not only made just for you. So personal, you know, things are very important, and none of the apps will take this in. **Participant 4, focus group 1**

And also, the things they tell you to do, they might not be, they might be suitable to some sisters. They might not be suitable to me, I'm a heart patient. I have had open heart surgery. I've got high blood pressure. I have ulcers, so you know, there's so many things I'm carrying along. So, if they [videos] say, tell me to eat something, I can't do that. **Participant 1, focus group 5**

I think just to add to that, the cultural diversity that we have is not always reflected in the results and the information that is provided, and so, you know, if you're treating me like a five foot nothing White woman, and I'm six foot and I'm Black, it's really different to the way that you would choose to treat me. **Participant 3, focus group 1**

**F1:** So, the next question is, how do you think digital services would help with your knowledge of diabetes?
**P5:** If they were tailored, then I'm sure it would help. But I think it can't help but be general so…
**P3:** Yeah, I agree with that.

[...]
**P7:** Because that's what I've found, a lot of the information seems to assume that if you are diabetic, you are overweight [inaudible chat] which is causing problems for me, because I'm not overweight. **Focus group 4**

**P2:** Most of what I like, know, is not from, 'it has this amount of carbs in this,' it's just I know that I've eaten a pizza before, so I know how much insulin to give over what time period.
[...]
**P3:** But like Participant 2 said, you do get used to what's in the food that you're eating. So, then you can administer your insulin according to what you're eating and, you know, sort of stuff to avoid if that's what causes your sugar levels to spike. So yeah, I'm not really a digital person. **Focus group 3**

**Accessibility**

**P7:** No, I don't agree with putting everything on an app. Certain things, yes, but medically, I don't think it would work properly, because it's too… Just a broad question mark. If you don't answer any of them questions, it doesn't affect you. Like I've had plenty of [inaudible 0:04:55] when I've actually phoned up or tried to make an appointment, and it says, "What's wrong?" You know, there's nothing on there that actually describes what's wrong with me. And then it says at the bottom, "Fill in what is…" And I'm not any good at writing or anything like that, so I'd rather be able to speak face-to-face or even over the phone with someone, explain what's wrong with me, and they can say, "Well, come in here," or, "Get to the hospital." Whereas an app will go, "Well, you don't reach our criteria, so…"
**F2:** Is it okay, [P7], just 'cos we won't have it on tape, that the reason you're not keen on the writing stuff is –
**P7:** I'm dyslexic.
**F2:** Dyslexic, thank you, just so we've got that, so we can make that clear. 'Cos other people will be in the same situation as well. **Focus group 1**

**P5:** Maybe I can say – only just to highlight the fact that we're obviously communicating in English and we do it fairly well [laughter] really, but I can only imagine how that can be another barrier if English is not your first language. The language used is very complex, and, you know, it looks like you have to have a degree to understand the content on some apps.
**P7:** Somebody English would have to have a degree to understand it. Some of the words come up and you think, "What?"
**P4:** It does, yes. **Focus group 1**

And sometimes when you've got a lot of apps on your phone, it can be overwhelming. And also, you know, digital literacy then. Sometimes, you know, I feel, oh, what is this? I don't know how to use it or, you know, what do I need to do? And then I just, I just leave it. I won't go any further with it because I just, I know, I just think, okay, let me just keep it simple, you know, try and eat the right food and, you know, maybe not bother with the apps apart from the Libre one, which is okay-ish. **Participant 1, focus group 3**

**P7:** So you've got to have an up-to-date phone that will actually accept that app?
**P4:** Yeah, it's got to have the NFC, like the same thing that you use for paying with the phone. So, it's got to have the NFC and that's pretty much all.
**P7:** It's not cheap.
**P4:** Well, you can get a phone like this probably for like eighty quid, I would say.
**P7:** Yeah, but if you're on benefits, you haven't got eighty quid. **Focus group 1**

Or of course, if somebody was to show you how to do something on a laptop or a computer, that's all very well, but if you then get home and you find, "Oh actually, I haven't got a fast enough phone or a fast enough laptop," well, it's a bit pointless really. **Participant 1, focus group 1**

It could be bilingual as well, you know. Different languages, you know, it can be different languages as well. Because some people, they cannot understand English. Some people speak their mother tongue like Urdu, Punjabi, Bengali, whichever language. **Participant 3, focus group 2**

I'm not…I can read and write, but...when I get a text, I have to go over and over and over, and to write a text…"I'll see you in an hour" or "Are we going here" or- it takes me probably 10 minutes to do it. I send the text and by the time I've put the phone down, they've replied to me and I think that, you know, I'm just a little bit slow and obviously, you know, that's why if I sit and talk to someone, I take it in and when things are important, they stick there, don't they? **Participant 6, focus group 4**

'Cos I think everything like that just needs to be made user-friendly, so more people can understand it rather than look at it and just go blank, which is what I do with a lot of stuff. **Participant 7, focus group 1**

Yeah, just about the digital literacy. I think Participant 3, you know, mentioned, I think, yeah, you know, it is good to use it. I say, you know, if something comes out, try it and see, because in some ways you can kind of get left behind a little bit. So, you know, but they maybe need to make it more accessible so that, you know, everybody can use it, make it very simple in the sense that not because people are silly, but, you know, to make, for it to be more inclusive then, you know, for everybody to be able to use it. Because I think there's that fear as well, even myself sometimes I think, 'oh, you know, do I want to be using that?' Or you press a button, you think, you know, you're going to share your business with everybody. You don't really know what you're doing. So, I think that could be quite helpful, just kind of simplifying it really. **Participant 1, focus group 3**

Yeah, 'cos Participant 2, you were saying about – and I've used some of those apps where you can look at the calorific value. It's a bit of an eye opener, isn't it? But you can only go so far before they then go, "Oh, you need to subscribe to this. You need to sign up for it. And by the way, it's going to cost you £25 a month," or whatever. You know, you're thinking, no, I don't think I'll do that, you know. So, it's very limited. **Participant 1, focus group 1**

**P3:** But like for us, people who don't know how to use it, it's very, very hard.
**P4:** The thing is that everything's online. Anything and everything is online, you know. If you go to the doctor or you go anywhere, online, online, online, but if you don't know anything about online, what can you do?
**P6:** There's a lot of people that are not online people though.
**P1:** Yeah, some people don't have internet at home, can't afford it.
**P4:** You're stuck then, aren't you, then you're relying on other people. **Focus group 2**

**P1:** Well, like I said before, erm particularly elderly people, they can't all use the digital things like the laptops and the phones and things like that, I know my husband can't.
**P7:** A lot of them don't actually have them.
**P1:** No.

| |
|---|
| **P7:** When they do, they're very reluctant to use and they'll just use them for one basic thing and then scared to do anything else. **Focus group 4** |
| **Reliance on family member support** |
| And the other thing is, you know, like we were saying that – what do you call it, us, we're not very familiar with apps and everything, yeah? And what do you call it, you want to know more information, you know, how everything is, right? But one way that – you know, when we rely on the kids, right, kids are too busy with their own personal lives. They've got time, they will do it for you. If they haven't got time, "Okay Mum, okay, I'll do it in a minute." **Participant 3, focus group 2**<br><br>**P6:** Because my, sorry, my children, they can explain [to] me. Sorry, they can explain [to] me. If it's any digital, they can watch wherever they want. Computer or a phone, they will tell you. They will show me how to use it. I don't know how to use computer yet.<br>**T2:** She would still be relying on someone to [help her] use her. **Focus group 5**<br><br>Well, I think I've been fortunate because I mean, my wife's a retired GP, so she accesses all – I have six monthly blood tests, not just testing diabetes, but testing a range of other things, and, you know, when you access this online – and I let [name of wife] do it. Really, I should learn to do it as well. So, you know, I've just been very fortunate that the surgery monitors me quite well. **Participant 6, focus group 1**<br><br>Sometimes there's too many links, "Click this link for that, this link for that, this link for that." How do I know, 'cos it's not been explained to me in the first place, you know, what I'm actually looking for. My daughter does – I mean, if I get a new medication or – she looks up a diagnosis or something for myself or whatever. She will look it up then, but I've got no practical way – I don't know how to do that. I'm a person that – if you give me a sheet and it says, "Do this, do that," it means nothing to me, but if you stand there and go, "If you do this and you do that and you do that," I will take it in. But if you give me a sheet and say, "Steps one to ten," by two, I've lost interest. **Participant 7, focus group 1**<br><br>**F1:** So, what is it that you would find hard if like you spoke to a GP and he said, "Go online and look at how to manage your diabetes?"<br>**P4:** I'd go and see my daughter, but for me, I can't.<br>**F1:**What is it that you would find difficult?<br>**P4:** To search myself, anything like that.<br>**F1:** Is it knowing what to search for or how to search?<br>**P4:** I'm going to wrong app and – because I've not got much knowledge – when I was in school, we had no IT, no nothing. **Focus group 2** |
| **Hearing other people's experiences important (friends, family)** |
| I think from my experience, I found that talking was more better, coming from another person, like a group of people, talking about their own experience. Like everything's not good for everyone. Whatever's good for me, it might be not good for the next person. And on top of that, you know your own body, what you're putting in. So like I mean, this lady called [Name] who delivers this healthy eating programme, and the way she comes out with the words and explaining, I found it quite good. And then she asked, "Tell us if you've got any questions, ask," and everything. I mean, there's a few groups of people and some people are like, [inaudible 0:25:31] to ask a question again and again, but other people still learn from that as well. **Participant 4, focus group 2** |

| |
|---|
| I think for me, yeah, the digital side of it. I don't know, it quite sort of scares me because I like this sort of support, you know, talking to another diabetic. I don't think you can get better than that. And I think sometimes, yeah, I can feel sort of quite isolated. And it, like, it's not another person, it's just an app. Like, and I, you know, I don't want to be like afraid of living with diabetes or I don't want to, you know, I don't want to feel isolated. And I think when these sorts of new digital stuff is coming out, it's like, 'oh, something else I've got to manage on my own.' Like, I like to be sort of quite open. You know, I've got it. There's nothing I can do about it. And I think sometimes like to make other people aware is the best sort of support that you can get because, you know, if you're in a crisis, an app's not going to save you. Do you know what you mean? If you know, by picking up the phone and talking to another sort of diabetic or you know, yeah, that's the best sort of support. So, I haven't sort of downloaded many apps, but yeah, I have got the Carbs and Cals, but I don't like use that strictly. **Participant 3, focus group 3**<br><br>**P1:** So, both are, but in person it does help people, bring people out. Gather them. Only here, the sisters are here, only one man, but at least we're here. We see each other, talk to each other. It brings people out of the house.<br>**P4:** Like me now, I learn from brother saying that he hasn't eaten roti, like chapati, for six months. Then I can't do without chapati because I can eat anything else. But as long as I eat one chapati, I'm fine [laughs]. But like, even I go on a holiday, I'm looking for my food. You know. **Focus group 5**<br><br>**P3:** I think, I belong to a couple of groups on like social media and I find them really, really helpful. Yeah, again, because you know, people are really talking about their experience and they're giving you advice based on their own experience. So again, I'd rather sort of talk about it than just look at it on a screen.<br>**F1:** Yeah. And what if, Participant 3, like an app, for example, had a social support aspect within it, you know, whether that be a forum or some sort of messaging component or buddying scheme, like is that sort of thing, would that be interesting to you or not?<br>**P3:** Yeah, yeah, yeah, it would be much better. Yeah, I would, I would take part in that for sure. **Focus group 3**<br><br>I guess it's nice to have the option. Do you know what I mean? Because like you say, you know, sometimes at 3 o'clock in the morning when your sugar level is going down really fast, there's not really anybody to talk to. So, I think that's the time that I would probably use the digital side of it. You know, if I was on my own, there was nobody around and it was in the middle of the night, that's when I'd be more inclined to use the digital stuff. **Participant 3, focus group 3**<br><br>I was just going to say about, I think you asked earlier about, you know, how, what sort of support would you like to see? And I think something like this, if you was able to, you know, we talked about waking up in the night or, you know, really worried, being able to log on, even if the people were not from, I don't know, London or wherever, you know, regional, then, you know, where you could literally log on and see people and talk to them if you can't be with them physically, you know, even if it was on an app, you know, I think that would be a really good idea. You know, sort of a 24-hour thing where people maybe can get access to actually speak to other people if you're really, you know, feeling that you can't deal with it. Because there's times, even as a type 2, and I've been, what, it was eight years. Gosh, it can be difficult. It's not easy and it's hard when you haven't got anyone to talk to about it, who's going through the same thing. **Participant 1, focus group 3** |
| **Theme 3: Healthcare structures shaping digital engagement** |
| **Digital tools and healthcare working together** |
| **P4**: Yes, and the [cancer hospital] did have a specialist diabetes team, which said the technology – I'm not sure it works at the moment, but the technology is that your readings get transmitted to a team of your choosing.<br>**P3:** What, that does?<br>**P4:** Yeah. So, you can ring them up, talk about a problem. They just pull up your graphs and come up with a recommendation as to how to handle things. |

**P8**: See that's good use of technology [inaudible]. **Focus group 4**

But I think it's also easy access as well, because since I've got that app from the doctors, I've been more in contact with the nurse, the diabetic nurse, than I ever was before. I just think it's marvellous. **Participant 3, focus group 4**

**P1:** Some aspects of technology, I've found quite helpful, 'cos being reasonably recently diagnosed and having had that experience of never going to the doctor, I didn't understand a thing about NHS apps, but I actually found the NHS app was really useful, to be able to look up and see what your results are, and then actually look at them and go, "What the heck is that?" And you can then look at it, and I thought that was very helpful, to explain how that A1c, whatever it is test –
**P4:** HbA1c.
**P1:** Yeah, that one, that one [laughter], how that actually sort of works, you know. You sort of read up about it and you go, "Oh, I understand that now," and then you think, "Oh no, I don't really." But I mean, that was really useful, and I hadn't realised just how much information you can get from the NHS app. 'Cos I went in there the first time to see somebody, and he just sort of spouted at me, and he said, "Oh yeah, do this, do that," and I came out and I thought, "I've no idea what I'm meant to be doing. Why didn't he write that down for me?" And of course I realised then, when I looked at my NHS app, it was all on there. Nobody said that to me, mind you, so nobody explained to me how to use it. That's your problem, if nobody explains to you how to use it then you don't know. **Focus group 1**

**F1:** So, what do you think digital services for diabetes self-management are trying to achieve? So, what do you – why do you think they're making things digital? And why do you think – what do you think they intend to do when they do that?
**P3:** Time, time, they don't have time to go to the doctor every time.
**P1:** It's also faster. If you can rely on the digital media, it's faster to get information, because nowadays you ring up a doctor and first thing they say, "What do you want? He's on holiday two weeks, [can] give you appointment for two weeks." "I might not even be in this world in two weeks!" I said, "What are you talking about!" So, the trying to, like the lady said, save time, save national health money and bring people onto digital, create a digital mind. The liability is when people have suspicion about the information on the digital media platform. That is what they have to improve, get people to trust the medium. **Focus group 5**

I think understanding what is out there is probably. I think once we know that, then we can decide which apps are more appropriate. **Participant 8, focus group 4**

**Experience of healthcare is dependent on where you live**

It varies from one region to one GP in the same area. Yes, you can have the same area and can have one GP with a different policy to another GP – one will see you and one won't. **Participant 8, focus group 4**

**P3:** Yeah, I just feel an app that would get me appointments would be really useful [laughter].
**F1:** Is that appointments with your GP?
**P3:** GP or the – 'cos you do your annual – "you must have it in September". So you phone up, and they say, "The first appointment is in October." "Okay, but you sent me a letter to say it must be in September."
**P7:** That's because they send that to every other body, and they've already booked up. **Focus group 1**

| |
|---|
| One thing, these days, you can't get appointment with a doctor. And maybe keep ringing you 'in line, in line', and when you get through, all appointment goes, so difficult to get. **Participant 6, focus group 5** |
| **Suspicion of digital tools taking over healthcare** |
| I mean, surely the point is that this will be a support but in addition to face-to-face contact with the diabetic nurse? Or do you think it's going to take over? **Participant 6, focus group 1**<br><br>**P3:** Well, do you think in years to come that there won't be… you won't be going to go in to see a doctor?<br>[…]<br>**P8:** But I think the way that we're moving and the pace at which we're moving, that doctor will become an AI doctor. **Focus group 4**<br><br>I think the, well, as the ladies are saying, they're more comfortable in person, but like I said in the beginning, very soon you won't be seeing doctor in person. It'll be AI sitting on the other side. So, you'll have to get used to getting information, whatever you need from the computer, because that's the future. **Participant 1, focus group 5**<br><br>**P6:** No way. And I think you can be a bit suspicious when you think of what NHS trends might be in the next ten or twelve years, with more and more maybe American influence, and you wonder –<br>**P7:** Well, they're advising you, you've got to get insurance now.<br>**P6:** You wonder how accurate or impartial the information might be, or is it leaning towards supporting a particular product that they want you to use. And in fact – I won't say anything more, but my brother-in-law very nearly lost his job with a charity because he queried… a particular company, and he – they were arguing, the TV adverts, that a particular tablet was suitable for children, and it was only for adults. But it was sorted. They were actually – it was a blatant lie. **Focus group 1**<br><br>In a lot of GP practices, you're not able to go in and see a doctor face-to-face. So, if we're moving with the times, that's where I can see that gradually technology taking over the personal touch that we've experienced. Which is lacking even now. **Participant 8, focus group 4** |
| **Value of F2F relationships** |
| **F1:** Yeah, it sounds like you've got quite a lot in your community, that face-to-face support?<br>**P3:** Because the thing is, you know, when somebody is talking, you know, even if it's not in words, right, you know, your face impressions, you get all the message. So, with online, yes, yeah, they give you this and that, but you know when you're – you don't need to say in words, but, you know, your face is enough.<br>**P1:** You can better understand, that's what she's trying to say. **Focus group 2**<br><br>**P8**: I personally would prefer it face-to-face […] Because I mean, they can actually, if you're talking about something that's happening, they can physically see your body?<br>**P2:** Your body language tells lots of things about you. **Focus group 4**<br><br>**P2:** I'm just thinking about, sorry… erm what you gain from having face-to-face and that what I was saying before about actually seeing somebody's expressions and you might not be able to portray that over the phone. But if they actually saw you, they'd realise that you wasn't well and think that's any [inaudible]. |

| |
|---|
| **P8:** So that choice where you, you know, you get the option of face-to-face, telephone, e-mail, you know, you choose. **Focus group 4**<br><br>But at the moment, because we have a different generation, you know, I'm plus 60, I'm not going to tell you my age [laughter]. But the younger generation, they will have no problem. Because they are phone users a lot. They can tell whether this source of information is honest, good, reliable, and they will take it. We have difficulty sussing that out and that's why we like to see diabetic nurse or the doctor for our own peace of mind. That's what I think. [Group agrees]. **Participant 1, focus group 5**<br><br>Well, they can do health checks and reviews, you know, like programmes in the centres as well, [Names of Community Centres], you know, when they're doing cholesterol and diabetes checks, and they give you advice as well face-to-face, I think that's more better. **Participant 6, focus group 2**<br><br>**P4:** I myself feel better to go to the doctors and [see them] face-to-face.<br>**P6:** My doctor, she's so good. Every time they call me, check-up. And nurse is so good. And she explained [to] me so nice what to eat, what not to eat, what time, and that's very nice. They keep checking us. **Focus group 5** |
| **Lack of support from HCPs** |
| Sometimes there's too many links, "Click this link for that, this link for that, this link for that." How do I know, 'cos it's not been explained to me in the first place, you know, what I'm actually looking for. My daughter does – I mean, if I get a new medication or – she looks up a diagnosis or something for myself or whatever. She will look it up then, but I've got no practical way – I don't know how to do that. I'm a person that – if you give me a sheet and it says, "Do this, do that," it means nothing to me, but if you stand there and go, "If you do this and you do that and you do that," I will take it in. But if you give me a sheet and say, "Steps one to ten," by two, I've lost interest. **Participant 7, focus group 1**<br><br>**P1:** And if you're going to use that app, then you need some real help and education on how to use it to the best of – you know, to benefit you, you know. Someone like [P7] here would need somebody to sit down and really look at it with you, and say, "Well, this is how you do it and this is where you go." Because if you just get shoved into an app –<br>**P6:** And that help to be available in two or three or four months or a year's time.<br>**P7:** Well, as I say, you get used to something and then all of a sudden they stop it, or they change it. **Focus group 1**<br><br>**P3:** I think most other people, they don't know how to use the app, to tell you the truth. Because you know, I think [inaudible 0:13:13] is a young one, if they give you, you know, training, how to use the app and then all that, I think they will get more better. Because as many people –<br>**P6:** They're not into all that.<br>**P3:** Yeah, so if they could give them training on how to – I think they will be okay.<br>**P1:** Then you'd be able to log into it and all that.<br>**F1:** And where could that training happen? Do you foresee that happening when you go for your healthcare appointment, or is that somewhere else?<br>**P1:** The healthcare one, they won't tell us, they only just check our bloods, that's all they do. The doctors don't tell you what to do, you have to go to the hospital, or –<br>**P6:** They say go on a link thing.<br>**P1:** Yeah, go on a link, "this is the link", yeah. **Focus group 2** |

**P8**: I think they've introduced technology, but they haven't educated the people to use the tools that they've brought in and as erm Participant 5 has said, it's confusing with all these different apps coming in. Some are linked to your hospital, some apps are linked to your GP, others are independent and they're all trying to do the same thing. So, if they were integrated into one single sort of portal that would help everyone. Plus, in fact, a lot of people are not digital erm...
**P3**: …savvy
**P8**: …savvy, yeah, and they will struggle and yeah, you might get help from your children or your siblings, but I erm, think it's just been, 'Here you go get on with it.' I think that's the wrong way to approach it. **Focus group 4**

**P4:** I think learning how to use the thing in the first place is the difficult thing.
**P8:** It's education, I think. You know, you need to be shown basically how to go through that process.
**P4:** And there are features that you don't use frequently so you forget that they're even there. And, you know, someone says, oh, just go to the homepage and find it on there and you are so used to just going into the prescriptions bit, you forget that there is the way of seeing your test results and so on. So, it's a matter of being shown. And then nowadays, they'll probably say there's a video on YouTube. But it's still better to be shown… **Focus group 4**

**F1:** How did you find out about the NHS app?
**P1:** 'Cos I think my husband said to me, he said, "You've got an NHS app, look it up and find out what your blood tests are."
**F1:** So it wasn't a healthcare professional telling you?
**P1:** No, none whatsoever. And then I hadn't realised how good the NHS app is in some respects. I'm quite a [when my husband said to me? 0:18:04], "Oh, why don't you look it up?" You know, and then you go, "Oh, I didn't know I could do that," and then [laughter] – a good few people I've been talking to. But the NHS don't tell you and the doctors don't. They just assume you know what you're looking for. **Focus group 1**

I'm confident in using it but it's just not knowing everything. It's the understanding what's available to see what's useful and what's not useful. I think it's educating people on the availability and then letting them decide what way they want to interact with technology. **Participant 8, focus group 4**

I find digital tools – uh, I guess at my age, I have to master this simple digital tool before I can go any further. In the case is only of recent that these things have been introduced to people my age. […] You know, but there isn't much out there for all the people to learn about digital. **Participant 3, focus group 6**

**P2:** Of course, we need them to show us how to use it, what are the things to look for, what are the recordings and what the- What's the interpretations of the data? […]
**P1:** Yeah, we can go in a group. Somebody is at least. Maybe we have them show us, taking us step by step, step by step. Because me, if you want to teach me something, I like to visualise it, show me. **Focus group 6**
